# To TTV or Not to TTV? A Combined Urinary Biomarker Approach in Kidney Transplant Recipients Using Torque Teno Virus, Cell-Free DNA, and BK Polyomavirus Measurements

**DOI:** 10.64898/2026.10.13.26365472

**Authors:** Yazan Alhadid, Max Mamroth, Zhiwei Wang, Jacob Castaneda, Abhishek Chadha, Alexandre Vlassov, Anna K. Chikova

## Abstract

**Background:** Noninvasive biomarkers capable of detecting kidney allograft injury and distinguishing rejection from infection remain an important clinical need. Torque teno virus (TTV) reflects host immune status, but its utility in urine has not been well established. We evaluated urinary TTV, total cell-free DNA (cfDNA), cfDNA fragmentation, and BK polyomavirus (BKV) as potential biomarkers of kidney transplant rejection and BK polyomavirus–associated nephropathy (BKPyVAN).

**Methods:** Urine samples from 334 kidney transplant recipients were analyzed in association with kidney biopsies performed within 30 days of sample collection. TTV and cfDNA were measured by multiplex quantitative PCR (qPCR), with a subset analyzed by digital PCR (dPCR), and BKV was quantified by qPCR. Associations with biopsy results were evaluated using group comparisons and nominal logistic regression. Changes associated with outcomes of consecutive biopsies were assessed by matched pairs analysis.

**Results:** Among 544 unique urine samples, the urinary TTV/cfDNA ratio was significantly lower in rejection than in non-rejection samples (P<0.0001). Logistic regression adjusted for demographic factors confirmed an inverse association between the qPCR-derived TTV/cfDNA metric and rejection (adjusted odds ratio per delta Cq, 0.91; 95% CI, 0.88–0.94; P<0.00001). Significant differences were observed for all types of rejection. Association was confirmed by dPCR: higher TTV was associated with lower rejection odds, whereas higher cfDNA was associated with greater rejection odds. A nominal logistic model incorporating dPCR measurements achieved an AUROC of 0.76 (95% CI, 0.69–0.82). Matched pairs analysis showed a significant decrease in TTV/cfDNA when rejection developed and an increase following recovery from rejection, with no significant changes when biopsy status remained unchanged. BKV levels were strongly associated with BKPyVAN. No correlation between TTV and BKV was observed. Nominal logistic regression integrating TTV, cfDNA, and BKV measurements yielded AUROCs of 0.75, 0.97, and 0.73 for rejection, BKPyVAN, and non-rejection, respectively.

**Conclusions:** A reduced urinary TTV/cfDNA ratio is associated with biopsy-defined kidney transplant rejection, and changes in the ratio correlate with transitions in biopsy results. Combined TTV, cfDNA, and BKV measurements showed differential associations with rejection and BKPyVAN, supporting further evaluation as complementary urinary biomarkers.

## 1 Introduction

The long-term success of organ transplantation requires continued post-transplant monitoring to maintain graft function while balancing the risks of rejection, infection, drug toxicity, and other complications associated with prolonged immunosuppression. Rejection may develop before substantial changes in conventional measures of graft function become evident, creating a need for surveillance approaches capable of identifying allograft injury at an early and potentially reversible stage. Biopsy remains the diagnostic gold standard for organ transplant rejection. However, biopsy is invasive, carries risks of organ injury, and is not well suited for frequent longitudinal surveillance [1].

Considerable effort has therefore been directed toward development of non-invasive methods for post-transplant monitoring. Measurement of donor-derived cell-free DNA (dd-cfDNA) in blood has emerged as an important approach for detecting allograft injury and is increasingly incorporated into clinical management of transplant recipients. Nevertheless, currently available dd-cfDNA assays have some limitations. Elevated dd-cfDNA reflects graft injury but is not specific for rejection and may also occur with infection, ischemic injury, and other pathological processes. Diagnostic performance may also differ according to the type of rejection, with generally better performance reported for antibody-mediated rejection than for some forms of T-cell-mediated rejection (TCMR). In addition, cost, dependence on specialized testing platforms, and turnaround time may limit frequency and availability of this test in clinical practice [2]. Thus, there still is an unmet need for lower-cost informative biomarkers that may be suitable for longitudinal monitoring of transplant patients.

One actively investigated biomarker of functional immune status is torque teno virus (TTV). According to several reports, prevalence of TTV ranges from 65 to 100% across different human populations [3,4], and TTV levels have been associated with clinically relevant outcomes that correlate with the degree of immunosuppression [5–10]. Since TTV replication is generally greater under stronger immunosuppression, TTV viral load has been proposed as an indicator of the net state of immunosuppression in transplant recipients [10,11]. Multiple clinical studies have demonstrated that increased TTV titers are associated with an increased risk of viral infections [12–14] and tumorigenesis [15]; however, TTV has not been established as a causative agent of disease and is therefore classified as a non-pathogenic virus [16–18]. Thus, the potential clinical value of TTV is based primarily on its relationship with host immune status rather than on TTV-associated pathology itself. Two recently concluded clinical trials, TTV-GUIDE IT (EU CT-Number: 2022-500024-30-00) in kidney transplant recipients [7] and VIGILung (NCT04198506) in lung transplant recipients [19], are expected to further evaluate the utility of TTV monitoring for guiding immunosuppressive therapy.

Several studies have reported lower circulating TTV levels in patients experiencing rejection, consistent with the concept that lower TTV replication reflects relatively greater immune competence or insufficient immunosuppression. A recent meta-analysis of kidney transplant studies similarly found lower TTV viral loads among recipients who developed acute rejection [20]. However, clinical evidence supporting the feasibility of measuring TTV levels in blood to detect active rejection is limited.

Different levels of TTV are found not only in blood but also in various human samples. Though TTV replication is largely dependent on bone marrow and white blood cells [21–24], detectable levels of the virus have also been identified in saliva and urine [17,25–31]. Here, we tested the hypothesis that TTV levels in the kidney and urinary tract may decrease due to inflammation and lymphocyte infiltration during kidney rejection, and these changes may be detectable in urine even when they are not detectable in the blood of transplant recipients. To test this hypothesis, urine samples collected from 334 kidney transplant recipients were analyzed in this study (Table 1).

**Table 1.** Summary of the information for kidney transplant patients.

| Parameter | Patient data |  |
| --- | --- | --- |
| Age; years, median (quartiles) | 53 | (41 — 63) |
| Age; years, min — max (range) | 18 — 79 | (61) |
| Sex; male / female, n (%) | 217 / 117 | (65 / 35) |
| Individuals with at least one documented rejection event, n (%) | 151 | (45) |
| Race; n (%) |  |  |
| Asian | 13 | (3.9) |
| Black | 71 | (21.3) |
| White | 242 | (72.5) |
| Pacific Islander | 1 | (0.3) |
| Not provided | 7 | (2.0) |
| Number of biopsy reports per patient |  |  |
| Median (quartiles) | 1 | (1 — 2) |
| Min — Max (range) | 1 - 4 | (3) |

Urine-based biomarkers are particularly attractive for kidney transplant surveillance because urine is not only a non-invasively collected sample but collection also can be managed by the patients outside of clinical settings, potentially allowing a reduction in office visits. A variety of urinary biomarkers, including chemokines, cellular RNA signatures, proteins, and donor-derived or total DNA, have therefore been investigated as indicators of kidney allograft injury and rejection [32,33].

Urine is a challenging sample type because analyte concentrations can vary substantially as a result of both pathological processes and normal physiological factors, including fluid intake, dietary preferences, sex, and age. Appropriate standardization is therefore important for quantitative urinary biomarker analysis. One commonly used method for normalization of quantitative urinary tests is the measurement of urinary creatinine levels [34]. However, creatinine may not represent the optimal normalization strategy for DNA- or RNA-based assays. For this reason, total urinary cfDNA was selected as a second analyte for evaluation in this study.

Urinary cfDNA has been investigated in several previous studies, with particular emphasis on donor-derived cfDNA (dd-cfDNA) as a potential marker of kidney allograft injury and rejection [34–38]. While some studies have demonstrated associations between kidney transplant rejection and total cfDNA or dd-cfDNA levels in urine, others have reported no or only minimal associations [36].

This study evaluated total urinary TTV and total urinary cfDNA, the ratio of TTV to cfDNA levels and the fraction of longer DNA fragments in urine samples collected from kidney transplant recipients at different time points after transplantation. Follow-up analysis also included quantification of BK-virus (BKV) using commercially available assay. Data were evaluated for possible association with the results of kidney biopsies performed within 30 days from sample collection.

## 2 Materials and methods

### 2.1 Human samples used

Urine samples collected from kidney transplant patients were purchased from a human sample biobank collected by Verici Dx plc (Franklin, TN, USA). Samples collected by Verici Dx were part of the VDX 20-1001 study, which was approved by the Advarra Institutional Review Board (Pro00049177), and all participants provided written informed consent. Participants were enrolled in the study from March 2021 to January 2023.

Biopsy results reported for each test were summarizing reports originally provided by a certified clinical pathologist. Biopsy data provided by Verici Dx included the presence or absence of active rejection, with types of rejection defined following Banff classification in histopathological analysis. Results of SV40 staining were also provided. Additional available information included biopsy date, reason for biopsy (for-cause or protocol biopsy), transplant date, sample collection date, patient’s age, sex and race.

Samples were collected at various times post-transplant, while most available biopsy data were derived from standard protocol biopsies performed at 3 and 12 months post-transplant. Over 70% of all urine samples were collected on the day of biopsy, 80% of all samples were collected within 5 days before or after biopsy, and 20% of all samples were collected between 6 and 30 days before or after reported biopsy. If multiple samples for the same patient were available, only one sample was associated with each biopsy based on the nearest collection date. Samples collected over 30 days from the date of biopsy and samples associated with inconclusive histopathology were not included in this study. The study included biopsy-matched urine samples collected from 334 patients (Table 1).

Ethical sample collection and IRB compliance were assured by Verici Dx. Patients were deidentified with limited demographic and clinical information disclosed by Verici Dx. Patient identities were not shared with researchers and no direct access to patient information was provided for any member of the research team.

Information related to post-transplant patient treatment, immunosuppression method, infectious disease history or any other post-transplant dynamic in patient condition or management was not available. Only biopsy data and limited demographic information were provided for the purpose of the study.

Samples were transported on dry ice and stored at -80°C prior to DNA extraction.

### 2.2 DNA Extraction from urine

Urine samples used in this study were frozen after collection without any additives, pre-processing or pellet removal. Frozen urine samples were thawed and equilibrated to room temperature. Samples were gently mixed to fully solubilize any residual crystals. Urine was clarified by centrifugation at 2,000 × g for 20 min to pellet cellular debris and insoluble protein aggregates, supernatant was used for downstream processing. Extracellular vesicles (EV) and other particles remaining in clarified urine were concentrated using a modified Total Exosome Isolation Reagent (from urine) method (Thermo Fisher Scientific). The resulting pellet was collected by centrifugation at 10,000 x g for 1 hour when extracted from 1 mL of urine, and for larger volumes (3 – 8 mL of urine), a bucket rotor with centrifugation at 4,816 x g for 1 hour was used. DNA was extracted from the resulting pellet using EZ1&2 Virus Mini kit v2.0 (Qiagen^TM^).

For absolute quantification purposes, calculations performed during data analysis included adjustments for differences in sample volumes where applicable.

### 2.3 Real-time PCR

For quantification of TTV and cfDNA in urine, previously described qPCR conditions were used [39] with addition of a second internal control for detection of shorter cfDNA fragments. As described previously [39], TTV was detected by FAM fluorophore. Human beta globin (HBB) targeting assay hereafter identified as BGIC was detected by the JUN fluorophore. New primers and a probe labeled by the VIC fluorophore were added to the mix for quantification of shorter cfDNA fragments (SHORT control). All three markers were tested in single multiplex TaqMan reaction.

A BK virus (BKV) quantification qPCR test (Vi03453401_s1) and TrueMark Custom DNA control for BK virus (A50377) - a reference standard, were used as recommended by the manufacturer (Thermo Fisher Scientific).

All real-time PCR reactions were performed using QuantStudio 5Dx or QuantStudio 5 instruments (Thermo Fisher Scientific).

### 2.4 Digital PCR

All digital PCR reactions used the QIAcuity Probe PCR Master Mix (Qiagen) and the QIAcuity Nanoplate with 26k partitions per well (Qiagen) to run on the QIAcuity One, 5plex Device (Qiagen) as described previously [39] with one small modification: the SHORT control was added to the reaction mix with primers and probes concentration identical to those for BGIC. Considering the limited linear range of the digital PCR method, multiple dilutions of DNA were tested for the majority of samples. Digital PCR results were analyzed using Qiagen software. The concentration of TTV and cfDNA in stock solution was calculated using data from the sample dilution generating the most reliable data considering the established linear range for digital PCR method used. TTV and cfDNA quantifications were expressed as number of genomic copies per 1 mL of urine or as a ratio between selected markers.

### 2.5 Data Analysis

Statistical analysis was performed using JMP Pro software (JMP Statistical Discovery LLC). In all reported statistical analysis results, presented confidence intervals (CI) were 95% confidence intervals. For the area under receiver operating characteristic curves (AUROC) and precision recall curves (AUPRC) 95% confidence intervals were calculated by a bootstrap-based method using 2500 sampling runs, or, for model comparison, AUCROC, confidence intervals were calculated by DeLong method [40] using JMP Pro; and 95% confidence intervals for adjusted odds ratios were calculated by a profile likelihood-based method [41] by JMP Pro software. Two-sided p-values and confidence intervals are reported unless specified otherwise.

## 3 Results and Discussion

### 3.1 Analysis of total cfDNA and TTV in urine samples

The sample preparation method used in this study was designed for extraction of cfDNA and viral particles from extracellular vesicles present in urine samples.

Previously published studies analyzed the urine pellet, containing cells and cellular debris originating in the kidney and urinary tract [25, 42]. However, the urine pellet is a known inconsistent type of sample highly impacted by sex, age and other biological factors, and cfDNA and free viral particles present in urine are missed by this method. Measuring cfDNA and viruses in pellet-free urine requires extraction from a large volume of sample to generate sufficient yield, introducing technical challenges for routine extraction procedures. To address these difficulties, we established a process for cfDNA/viral extraction from cell-free urine samples. First, cellular pellet and other debris were removed from urine by low-speed centrifugation, next, extracellular vesicles and viral particles were concentrated using modified Total Exosome Isolation Reagent method, and the resulting pellet was used for total nucleic acid extraction. Though this method provided slightly lower concentrations of cfDNA, the yield of TTV was significantly higher in comparison to the yield obtained using commercially available kits suitable for cfDNA extraction from urine (Figure S1), suggesting that some cfDNA kits are less efficient in extraction of single stranded DNA, such as TTV. The method used in this study is expected to preferentially extract cfDNA from extracellular vesicles (EVs). Previous publications demonstrated significant differences in the composition of total cfDNA and the contents of EVs in urine, including differences in fragment size distribution and cfDNA origin [43], significant diversity in sizes and composition of EVs observed in urine under different conditions were also reported [44,45]. According to previous reports, cfDNA in urine EV was considered more stable and less impacted by collection and storage conditions.

Though the selected sample preparation method provides the capability for DNA extraction from large volumes of urine, initial proof-of-concept study used 1 mL of urine per sample with 40% of total extracted DNA analyzed in a single qPCR reaction.

The previously described qPCR-based TTV quantification assay [39] was modified for the purpose of combined quantification of TTV and total cfDNA in urine samples. Since cfDNA extracted from urine tends to be highly fragmented [46–49], an additional control for shorter DNA fragments was added to the previously optimized qPCR mix to ensure accurate quantification of total cfDNA in each sample.

Human beta-globin internal control (BGIC) was initially developed to quantify human genomic DNA in whole blood and plasma samples [39]. A SHORT control added to qPCR reactions for the purpose of urinary cfDNA analysis targets a different conserved region of the human genome and is designed to amplify shorter fragments. SHORT control primers are capable of detecting DNA fragments starting from 60 bp, and BGIC control primers only detect targets that are over 150 bp thus allowing to assess the level of DNA fragmentation in each sample with ΔCq between BGIC and SHORT controls (ΔCq (VIC-JUN)) representing the fraction of longer cfDNA fragments.

We hypothesized that ratio of TTV to total cfDNA as well as a fraction of longer cfDNA fragments in urine may be associated with kidney transplant status reflected in corresponding biopsy results. The Cq and ΔCq values were used to estimate concentration of each analyte in tested samples and the ratios among them. The ΔCq between fluorophores detecting the SHORT control and TTV (ΔCq(VIC-FAM)) reflects the ratio of TTV to total human DNA in each sample: a positive ΔCq is detected when TTV copy number is higher than the total amount of human cfDNA, and a negative ΔCq means TTV copy number is lower than cfDNA.

### 3.2 Initial case-control study with 281 urine samples

An initial study was designed to test new sample preparation and analysis methods, and to evaluate potential association of TTV and cfDNA levels in urine with kidney transplant rejection. The study included 281 urine samples from 158 kidney transplant patients. All included samples were collected within 30 days from biopsy conclusively detecting active rejection (164 samples) or no rejection (117 samples). Samples were selected from an available biobank to represent different types of rejection including for-cause and protocol biopsies performed at different time post-transplant, and to present a sufficiently diverse population of the patients based on patient’s sex, age and race (Table S1).

The qPCR results exported as cycle of quantification (Cq) values were analyzed for their individual or combined association with biopsy results; ΔCq(VIC-FAM) representing the ratio of TTV to total cfDNA was significantly reduced in the rejection group as evidenced by both t-test (p-value, 0.0026) and non-parametric test (p-value 0.0015) with the 281 analyzed samples (Figure 1A). The median difference between rejection and non-rejection group was 3.48 ΔCq, corresponding to an over 10-fold difference in TTV fraction between groups representing two different outcomes.

**Figure 1.**
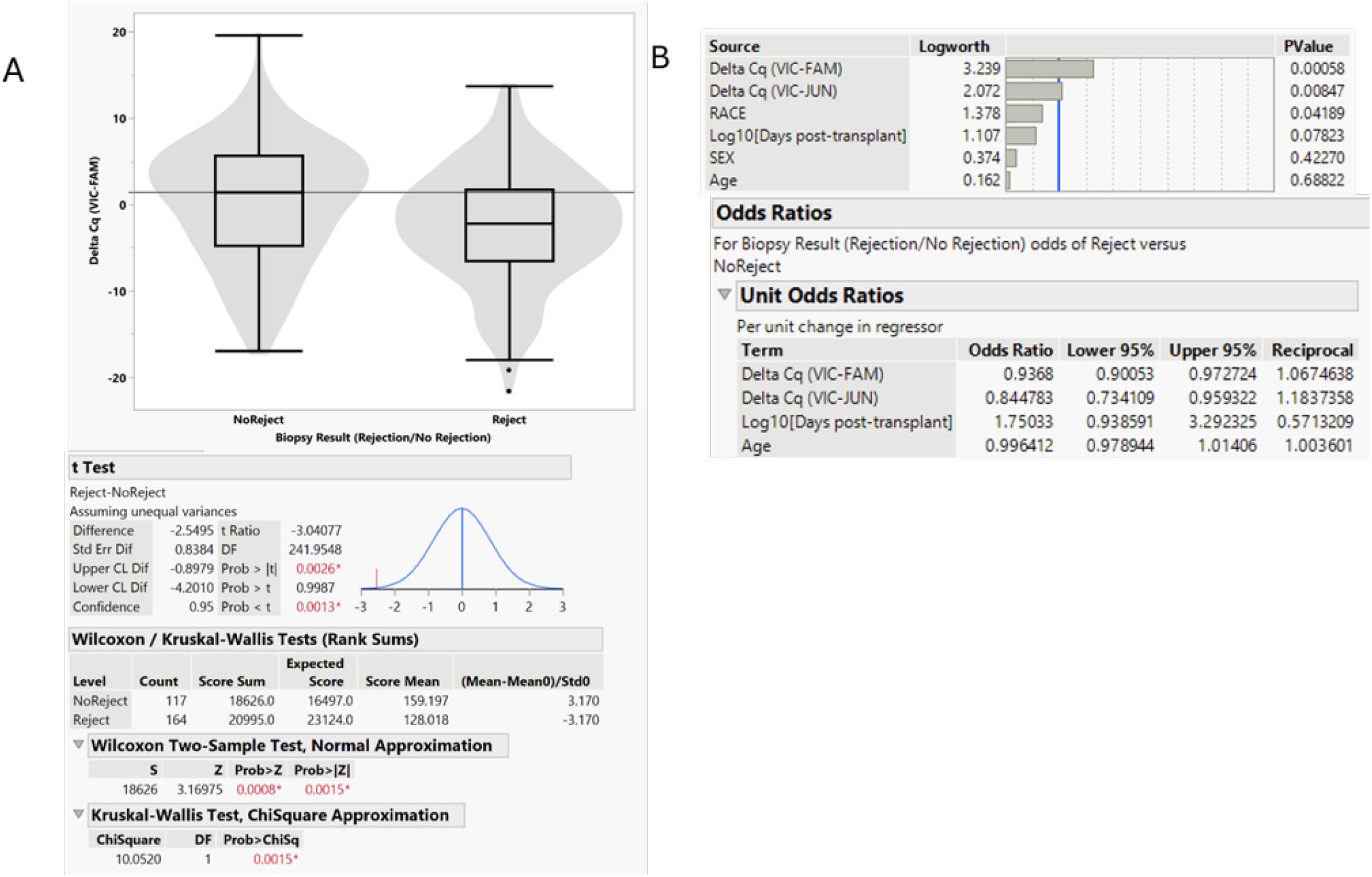
Results of initial case-control study including 164 rejection and 117 non-rejection samples. (A) Comparing ΔCq(VIC-FAM) in rejection and non-rejection groups using t-test and non-parametric tests; (B) results of nominal logistic regression adjusted by sex, race, age of the patient at time of transplant, and number of days between transplant and sample collection.

Logistic regression adjusted by the patient’s age, sex, race and the number of days between sample collection and transplant confirmed a significant association of ΔCq(VIC-FAM) with the risk of rejection (p-value, 0.00058), and adjusted per-unit odds ratio 0.94 (CI, 0.90 – 0.97) (Figure 1B) suggesting estimated 6% reduction in rejection odds with every ΔCq. Interestingly, though the TTV/cfDNA ratio was the most significant predictor based on this model, ΔCq(VIC-JUN) reflecting the fraction of longer fragments in total cfDNA was also significantly associated with rejection (p-value 0.0085), adjusted per unit odds ratio 0.83 (CI, 0.73 – 0.96) suggested possible changes in cfDNA fragmentation in urine as a result of active rejection. Though the t-test also showed a statistically significant difference in ΔCq(VIC-JUN) between rejection and non-rejection samples, the p-value generated in the t-test (0.043) was higher than in logistic regression, and the median difference in ΔCq(VIC-JUN) between rejection and non-rejection groups was only 0.16, suggesting that some of observed predictors are more significant after adjustment by other covariates.

Logistic regression analysis demonstrated independent association of TTV/cfDNA ratio and the level of cfDNA fragmentation with kidney rejection (Figure 1B).

For all qPCR reactions included in this study, a measurable level of at least one marker was detected. In cases when one of the analytes did not generate Cq value, an arbitrary Cq of 46 (total number of qPCR cycles plus 1) was assigned for the purpose of the delta Cq quantification.

Since in the initial case-control study nearly 13% of all tested urine samples demonstrated high or no Cq values for multiple markers indicating low DNA yield, 1 mL volume of urine used for DNA extraction in initial study was deemed insufficient for the purposes of the test.

To verify the association of TTV and cfDNA levels with rejection, a confirmatory study using an additional subset of urine samples was planned.

A follow-up study performed with additional samples used 3 – 6 mL of urine for the extraction with the intention to reduce sampling variability in downstream analysis. Samples demonstrating low DNA yield in initial study were retested with larger volume of urine if available.

### 3.3 Confirmatory study using additional 300 urine samples

To verify initial observations, additional 300 urine samples from 240 kidney transplant patients were tested. This study included retests for 37 samples analyzed in initial experiments (30 non-rejection and 7 rejection samples), comprising new DNA extraction from remaining urine volume and new qPCR analysis. The remaining 263 samples were unique for the confirmatory study and included all remaining samples available in the biobank and meeting inclusion criteria (no more than 30 days from conclusive biopsy). A summary of the patient/sample characteristics is displayed in Table S2.

The second dataset confirmed the association between a reduced ratio of TTV to total cfDNA and rejection observed in the initial set of 281 randomly selected samples. Both t-test and a non-parametric test comparing ΔCq(VIC-FAM) values between the rejection and non-rejection groups demonstrated p< 0.0001 (Figure S2A).

Logistic regression adjusted by race, sex, age of the patient at the time of transplant and number of days between transplant and sample collection was used to analyze confirmatory samples. Interestingly, ΔCq(VIC-FAM) was the only variable demonstrating a significant inverse association with rejection outcome among two subsets of samples (Figure S2B). The per-unit odds ratio for ΔCq(VIC-FAM) in confirmatory data subset reached 0.88 (CI, 0.84 – 0.92), which translates into an approximately 12% reduced odds of rejection with every ΔCq (Figure S2B). In contrast, ΔCq(VIC-JUN) did not show significant association with rejection in the second set of data.

To further assess whether limitations of qPCR technology impact the results of association analysis, additional tests using digital PCR method were performed. Though, as demonstrated before, qPCR is a reliable method providing large linear range of TTV measurements [39,50], the logarithmic nature of the Cq-based quantification may limit the magnitude of the changes that could be reliably measured by qPCR. Digital PCR (dPCR) offers a smaller dynamic range and may be more expensive, however, this technology is known for accuracy of measurement when specific copy number quantification is needed.

To check whether ability to detect small differences in DNA concentrations provided by dPCR method may be beneficial for kidney transplant monitoring, 258 residual DNA samples including 66 rejection and 192 non-rejection cases (Table S4) selected from the confirmatory subset were analyzed by dPCR in addition to qPCR. While relative measurements such as ΔCq and ΔΔCq were mainly used in qPCR data analysis, for digital PCR results, levels of TTV and cfDNA were analyzed as a number of genomic copies per 1 mL of urine. Calculations were adjusted with consideration of the double-stranded and diploid nature of the human genome. As TTV genome is presented by a single copy of single stranded DNA, no copy number adjustments were made for TTV quantification [51]. The calculated number of amplifiable genomic copies of TTV and cfDNA per 1 mL of urine were analyzed by using logistic regression and a partial least squares method in JMP Pro (Figure 2). For the purpose of this analysis dPCR results were converted into log10. For cases, when zero copies were detected for one of the analytes, for example, if a sample contained measurable cfDNA but no TTV or if only TTV was detected in the sample, an arbitrary number of 1 copy was assigned to the missing analyte for the purpose of the log10 conversion or ratio quantification.

**Figure 2.**
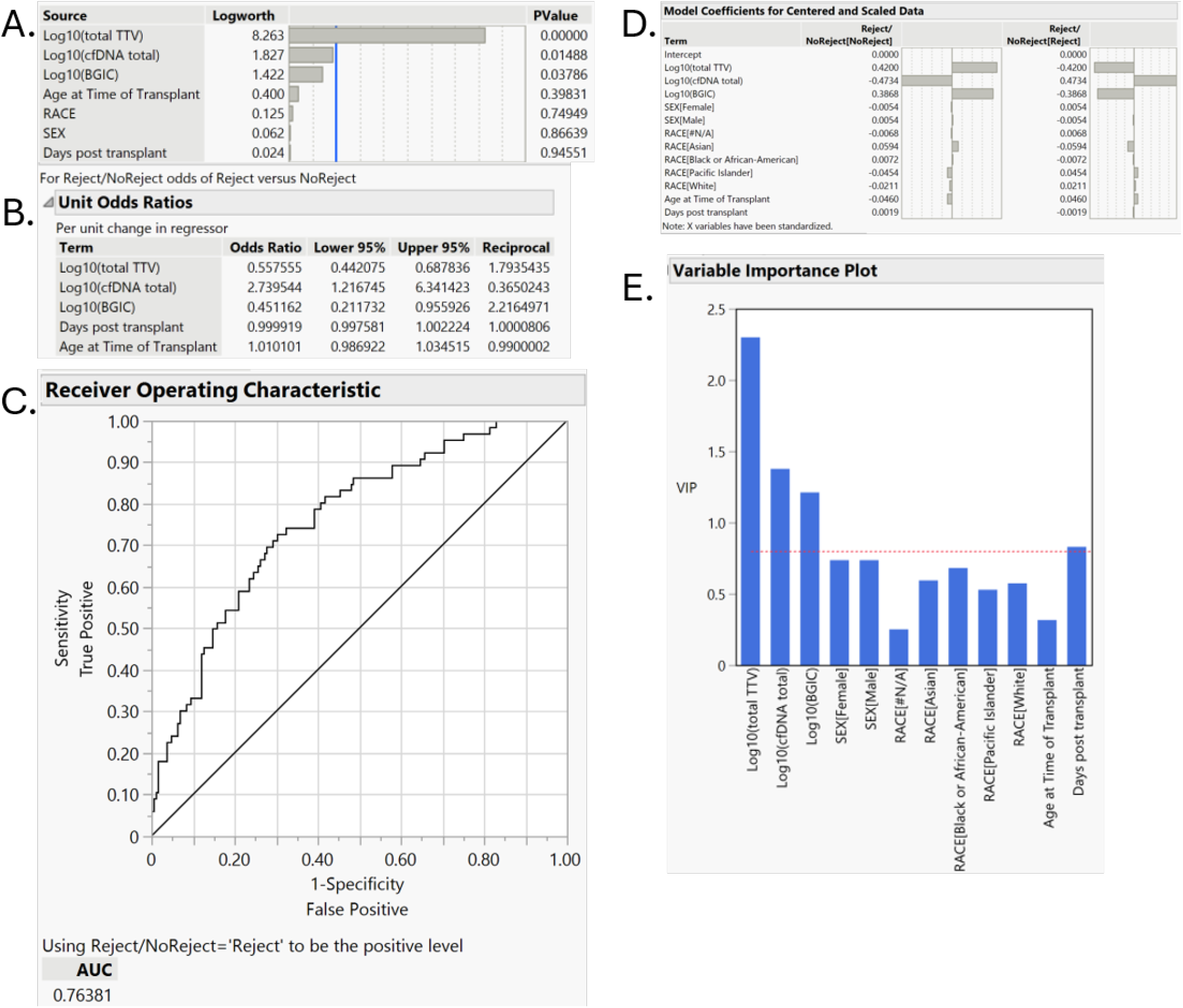
Results of dPCR analysis of 66 rejection and 192 non-rejection samples. (A) Effect summary of nominal logistic regression adjusted by sex, race, age of the patient at the time of transplant and number of days between transplant and sample collection. Model includes log10 of the copy number for TTV and cfDNA measured by SHORT and BGIC control assays and compares rejection and non-rejection biopsy. (B) Per-unit odds ratio (rejection/no-rejection) in nominal logistic model, (C) ROC curve for nominal logistic model. (D) Partial least squares model, normalized coefficients and (E) plot demonstrating relative importance of covariates in partial least squares model.

All three analytes measured by digital PCR demonstrated significant association with rejection (Figure 2A) with an odds ratio of 0.56 (CI, 0.44 – 0.69) per log10 of TTV and odds ratio of 2.74 (CI, 1.22 – 6.34) per log10 of total cfDNA copy number, suggesting an opposing association of TTV and cfDNA levels with kidney rejection (Figure 2B). Interestingly, quantification of longer DNA fragments showed a significant inverse association with rejection (p-value 0.038, adjusted odds ratios 0.45 (CI, 0.21 – 0.96) per log10 of BGIC copy number), confirming the observation from the initial case control study also showing association of increased cfDNA fragmentation with rejection. Despite the nearly negligible contribution of sex, race and age of the patient to the dPCR based logistic model, AUROC was 0.76 (CI 0.69 – 0.82) (Figure 2C). Partial least squares method of analysis demonstrated similar trends: the most significant association with rejection was shown for dPCR results, while age, sex and race of the patient did not demonstrate significant association with biopsy results (Figure 2D). Similarly to the logistic regression, TTV and cfDNA had opposite association with rejection, and the BGIC marker showed association opposite to total cfDNA quantification. Partial least squares analysis also demonstrated strong association of TTV with rejection odds followed by total cfDNA and BGIC. Number of days post-transplant was shown as fourth most important predictor in resulting model (Figure 2E).

The ratios between dPCR results for tested analytes were evaluated similarly to analysis used for qPCR data. A nominal logistic model including log10(TTV/cfDNA) and log10(BGIC/SHORT) adjusted by patient’s sex, race, age at the time of transplant and number of the days between sample collection and transplant showed the same AUROC 0.76 for relative quantification as for absolute quantification of each analyte. Interestingly, not adjusting the logistic model by patient information and only including dPCR results (log10(TTV/cfDNA), log10(BGIC/cfDNA) and total cfDNA), did not significantly reduce AUROC (0.75, CI, 0.68 – 0.81) and, moreover, nominal logistic fit using only log10(TTV/cfDNA) not adjusted by any other parameters provided AUROC 0.72 (CI, 0.65 – 0.78). Results demonstrated that the TTV/cfDNA ratio remains the main parameter associated with rejection when measured by dPCR, though combined analysis of TTV, total amount of cfDNA and the fraction of larger fragments may improve the discrimination ability of a predictive model. Differences between log10(TTV/cfDNA) and multi-factor models created with these data were insufficient to be defined significant under DeLong AUROC comparison analysis [40]. (Figure S3)

Directly comparing log10(TTV/cfDNA) in rejection and non-rejection groups showed a median difference of 1.5 log10(TTV/cfDNA) (Figure S4).

Overall, qPCR and dPCR results were in agreement, and both demonstrated strong association of TTV and cfDNA with biopsy results collectively confirming conclusions of the initial case-control study. Though digital PCR technology presents a challenge for viral quantification where large range of DNA input needs to be measured, combining results of qPCR and dPCR based analysis may be beneficial, in particular for samples falling in the range from -1 to +1 ΔCq in qPCR.

### 3.4 Combined analysis of 544 urine samples collected from 334 kidney transplant patients

Analysis of qPCR results for TTV and cfDNA quantification independently confirmed association with active rejection in two different sets of urine samples. Results were statistically significant in both data sets despite the relatively small number of samples in each. Combining data from both parts of the study could increase the power of statistical analysis and enable rejection type-based analysis. However, it was important to confirm that the two parts of the study were not systematically different.

To assess whether initial and confirmatory data are sufficiently comparable despite differences in the sample preparation procedure, two predictive models using the first set of 281 samples were generated. In addition to the qPCR data, this analysis also included time between transplant and sample collection, and patient age at the time of transplantation. A generalized linear model and boosted neural model were created using JMP Pro software. Models were analyzing rejection and non-rejection outcomes in the first set of data, and probability formulas resulting from these training data were applied to the data from 300 confirmatory samples (Table S3). Though AUROC for a generalized linear model was only 0.64 (CI, 0.57 - 0.71) applying the resulting formula to 300 confirmatory samples did not show reduced AUC value and, in contrast, demonstrated improved fit for confirmatory data with AUROC increased to 0.73 (CI, 0.67-0.78). Similarly, formula generated by neural model with initial case-control data with AUROC 0.68 (CI, 0.61 – 0.74) demonstrated slightly improved performance when applied to confirmatory data with AUROC 0.72 (CI, 0.65 - 0.77). (Table S3).

Similar analysis was performed for ABMR and TCMR separately. Despite a smaller number of samples available for each type of rejection in the first and second sets of data, a model generated with initial data for specific types of rejection showed comparable or better results in confirmatory samples. For example, a generalized linear model for comparison of ABMR and non-rejection samples showed AUROC 0.70 (CI 0.61 – 0.78), and the resulting formula applied to ABMR and non-rejection data from confirmatory samples demonstrated an AUROC 0.71 (CI 0.63 – 0.79). And for TCMR, a generalized linear model built with initial data had AUROC 0.69 (CI 0.57 – 0.75) with resulting probability formula generating AUROC 0.69 (CI 0.58 – 0.79) in the second study (Table S3).

These results suggested that though DNA extraction method was modified for a larger volume of urine in second study, data are sufficiently similar to be analyzed as a single set of the results. Combined analysis of two data subsets was expected to strengthen statistical analysis by increasing sample size for mathematical modelling and allowing additional data evaluation such as performing time-based analysis and, potentially, evaluation of less common biopsy results such as viral polyoma.

Combined analysis included qPCR results for only unique biological samples. For urine samples, which had to be retested, only retest data using higher sample input were included in the analysis. Table 2 shows summary statistics for 544 unique samples analyzed in the study. The number of samples used for each patient corresponds to the number of informative biopsies, results for which were provided. Thus, the number of samples tested per patient ranged from 1 to 4 (Table 1). Among 544 analyzed samples, 514 (94%) and 539 (99%) had detectable TTV and cfDNA respectively, and for 100% of tested samples either TTV or cfDNA or both were detected. For samples not generating Cq values for one of the markers, an arbitrary value of 46 was assigned for purpose of the ΔCq quantification and data analysis.

**Table 2.** Combined table of unique samples used for TTV and cfDNA quantification testing.

| Parameter | Sample Information<sup>a</sup> |  |
| --- | --- | --- |
| Age; years, median (quartiles) | 53 | (40 — 63) |
| Age; years, min — max (range) | 18 — 79 | (61) |
| Sex; male / female, n (%) | 352 / 192 | (64.7 / 35.3) |
| Race; n (%) |  |  |
| Asian | 26 | (4.8) |
| Black | 115 | (21.1) |
| White | 387 | (71.1) |
| Pacific Islander | 3 | (0.6) |
| Not provided | 13 | (2.4) |
| Biopsy Type, n (%) |  |  |
| Protocol | 391 | (71.9) |
| For-cause | 153 | (28.1) |
| Days post transplant, median (quartiles) | 109 | (88 — 285) |
| Days post transplant, min — max (range) | 4 — 685 | (681) |
| Biopsy results, n (%) |  |  |
| No Rejection | 312 | (57.4) |
| All Types of Rejection | 232 | (42.6) |
| Biopsy results for BKV associated polyoma, n (%) |  |  |
| BK Polyoma detected | 24 | (4.4) |
| No BK polyoma | 520 | (95.6) |
| Biopsy results, rejection type, n (%) |  |  |
| No rejection | 312 | (57.4) |
| ABMR | 68 | (12.5) |
| Borderline | 63 | (11.6) |
| Total TCMR including: | 51 | (9.4) |
| IA | 28 | (5.1) |
| IB | 13 | (2.4) |
| IB & IIA | 1 | (0.2) |
| IB & IIB | 4 | (0.7) |
| IIA | 2 | (0.4) |
| IIB | 3 | (0.55) |
| Mixed rejection including: | 50 | (9.2) |
| ABMR & IA | 10 | (1.8) |
| ABMR & IB | 4 | (0.7) |
| ABMR & IIA | 2 | (0.4) |
| ABMR & IIB | 2 | (0.4) |
| ABMR & III | 2 | (0.4) |
| ABMR, IB & IIA | 1 | (0.2) |
| borderline & ABMR | 29 | (5.3) |

<sup>a</sup> information is summarized for 544 samples obtained from 334 kidney transplant patients

Combined analysis of ΔCq(VIC-FAM) representing TTV/cfDNA ratio for 544 urine samples demonstrated significant difference between rejection and non-rejection groups (Figure 3A) shown by both two tailed t-test (p<0.0001) and non-parametric test (p<0.0001). Nominal logistic regression adjusted by race, sex, age of the patient at the time of transplant and number of days between transplant and sample collection showed strong reverse association of ΔCq(VIC-FAM) with rejection, with p<0.00001 and adjusted per unit odd ratio 0.91 (CI 0.88 – 0.94) (Figure 3B).

**Figure 3.**
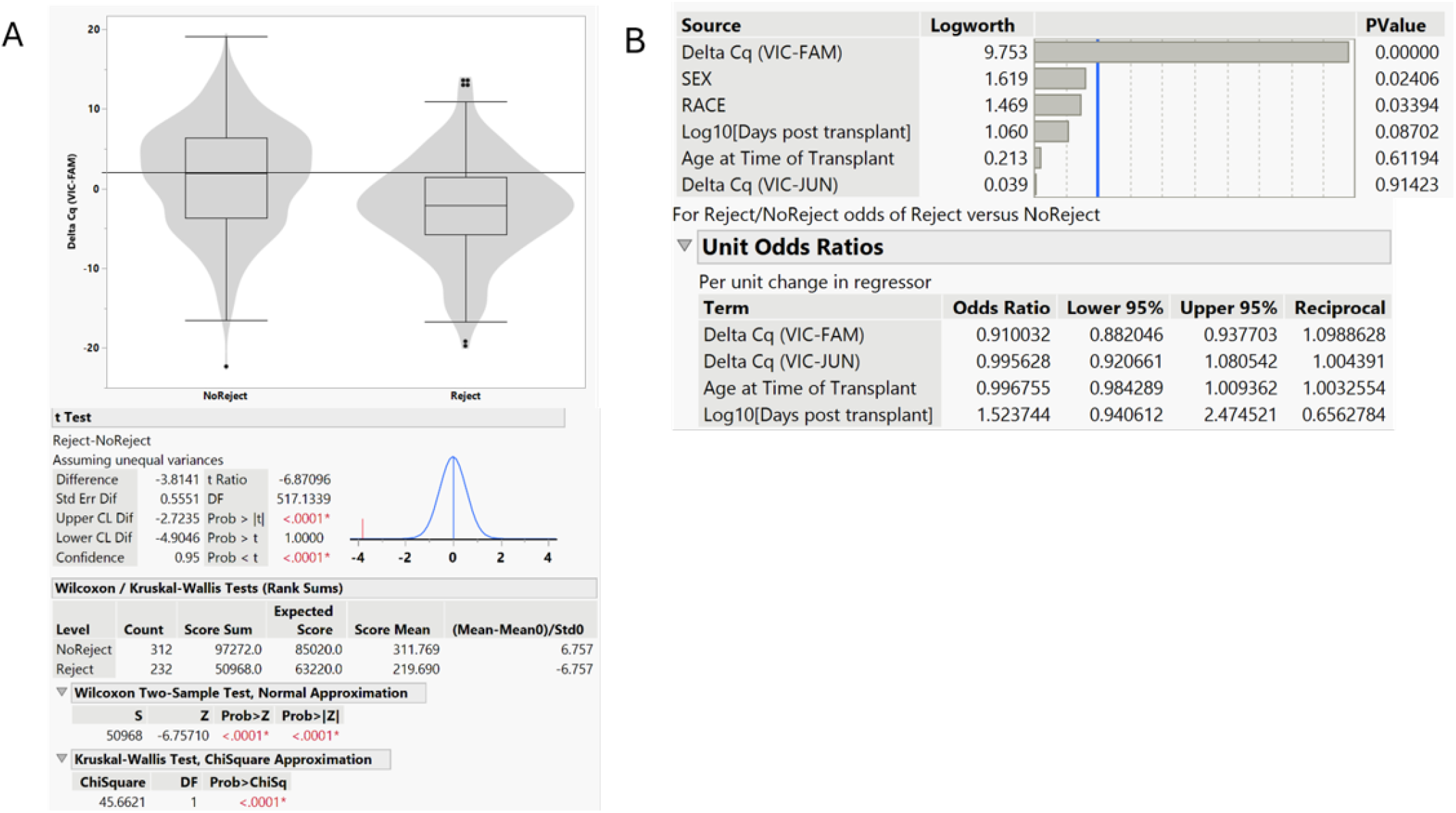
Combined analysis of qPCR results for 544 urine samples. (A) Comparing ΔCq(VIC-FAM) for 232 rejection and 312 non-rejection samples using t-test and non-parametric tests; and (B) results of nominal logistic regression adjusted by sex, race, age of the patient at time of transplant, and number of days between transplant and sample collection.

While adjustment of logistic regression models for demographic factors can account for covariation and potential confounding, inclusion of an excessive number of patient-specific characteristics may increase the risk of model overfitting and reduce the external validity of the identified predictors [52,53]. For this reason, an effort was made to minimize the number of covariates used in analysis to the most relevant factors and qPCR results to limit model complexity and reduce the risk of overfitting. The number of days post-transplant is a known factor impacting levels of TTV in patient’s blood, and thus may be an important confounding factor for TTV quantification results. Age of the patient at time of transplant is another known risk factor for transplant outcomes [54]. Though no significant correlation between age and rejection events was observed in this study, patient’s age at the time of transplant was also included into analysis.

A new nominal logistic model generated with qPCR results from 544 samples largely relied on urine biomarkers and included normalized DNA input values and ΔΔCq values combining analysis of TTV fraction with cfDNA fragmentation level in the sample. The model was adjusted by the number of days post-transplant and patient’s age. As expected, the new model showed strong association of ΔΔCq with rejection, p<0.00001 and adjusted odds ratio of 0.9 per each ΔΔCq (Figure 4A). The second and third most important predictors within the new model were total cfDNA and the number of days post-transplant with p-values <0.00001 and 0.02 respectively. AUROC of the resulting model was 0.72 (CI 0.67 – 0.76) (Figure 4B).

**Figure 4.**
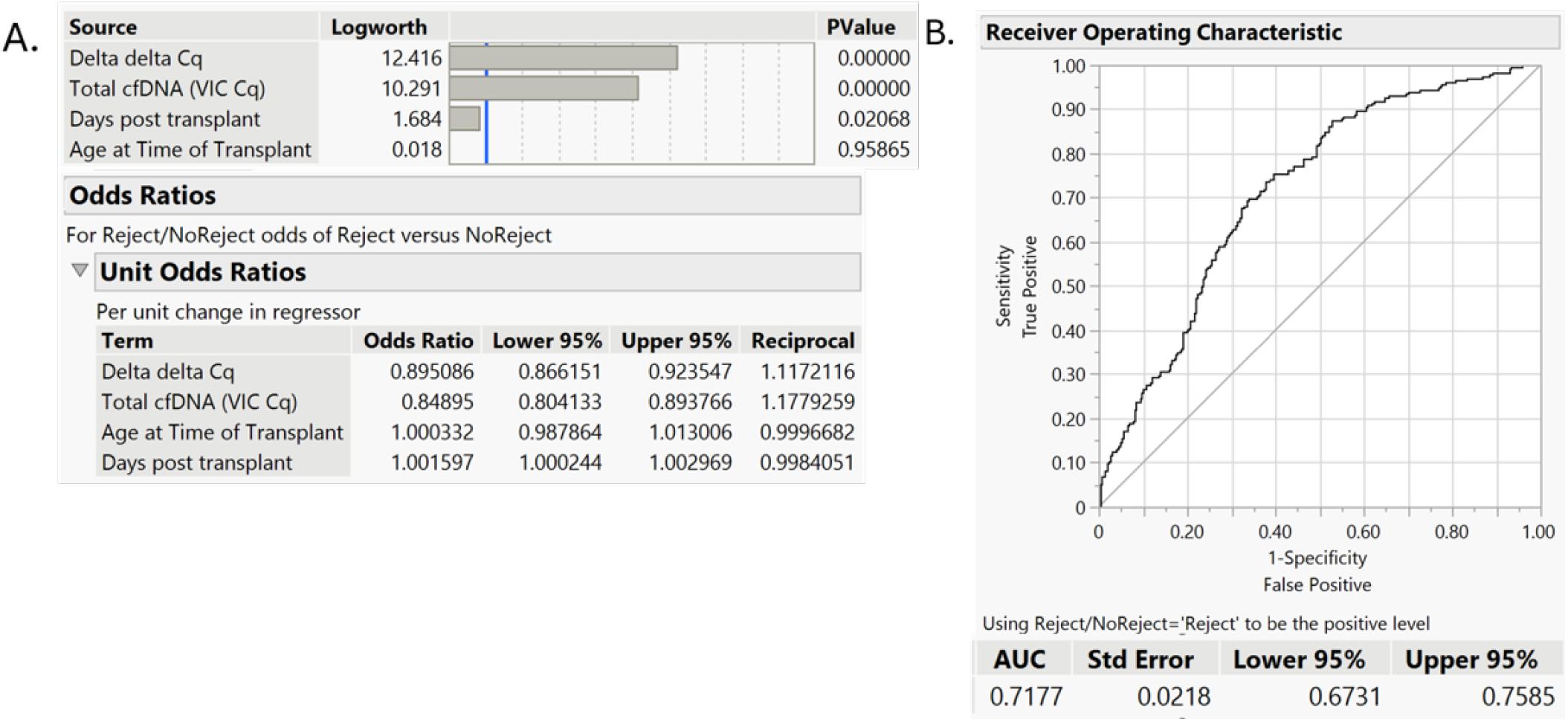
New nominal logistic model including ΔΔCq for qPCR data and total cfDNA quantification adjusted by age of the patient at time of transplant and number of days between transplant and sample collection. (A) Effect summary and per-unit odds ratios for nominal logistic regression based model, and (B) ROC curve and area under ROC curve with 95% confidence intervals calculated using the DeLong method.

Previously reported studies analyzing donor derived cfDNA in kidney transplant patients showed stronger association with specific types of rejection [55], and measuring total cfDNA in urine could be impacted by the similar phenomena. For this reason, analysis of ΔCq(VIC-FAM) among different biopsy outcomes was performed. Figure S5 shows distribution of ΔCq(VIC-FAM) for each type of rejection. Since different types of TCMR and mixed rejection were presented by only a few samples per each class, these types of rejection had to be grouped for the purpose of statistical analysis (Table 2). All rejection samples were separated into four groups: ABMR, TCMR, mixed rejection and borderline rejection (Table 2)

Different types of rejection were compared to non-rejection group (Figure 5A) by Dunnett’s method and non-parametric Dunn method for joint ranking. Statistically significant difference with each type of rejection was demonstrated by all tested methods with p-values ranging from <0.0001 to 0.0057 (Figure 5B). Interestingly, not only were all rejection types significantly different from control groups, every type of rejection demonstrated median ΔCq(VIC-FAM) below zero, while the median ΔCq for non-rejection group was above zero. Single sample t-test comparing each group with an arbitrary value of 0.5 showed that the distribution of the data for every type of rejection was significantly below this value, while ΔCq distribution for the non-rejection group was significantly higher than 0.5 suggesting a possibility to distinguish all types of rejection from non-rejection using TTV/cfDNA ratio in urine samples (Table S5).

**Figure 5.**
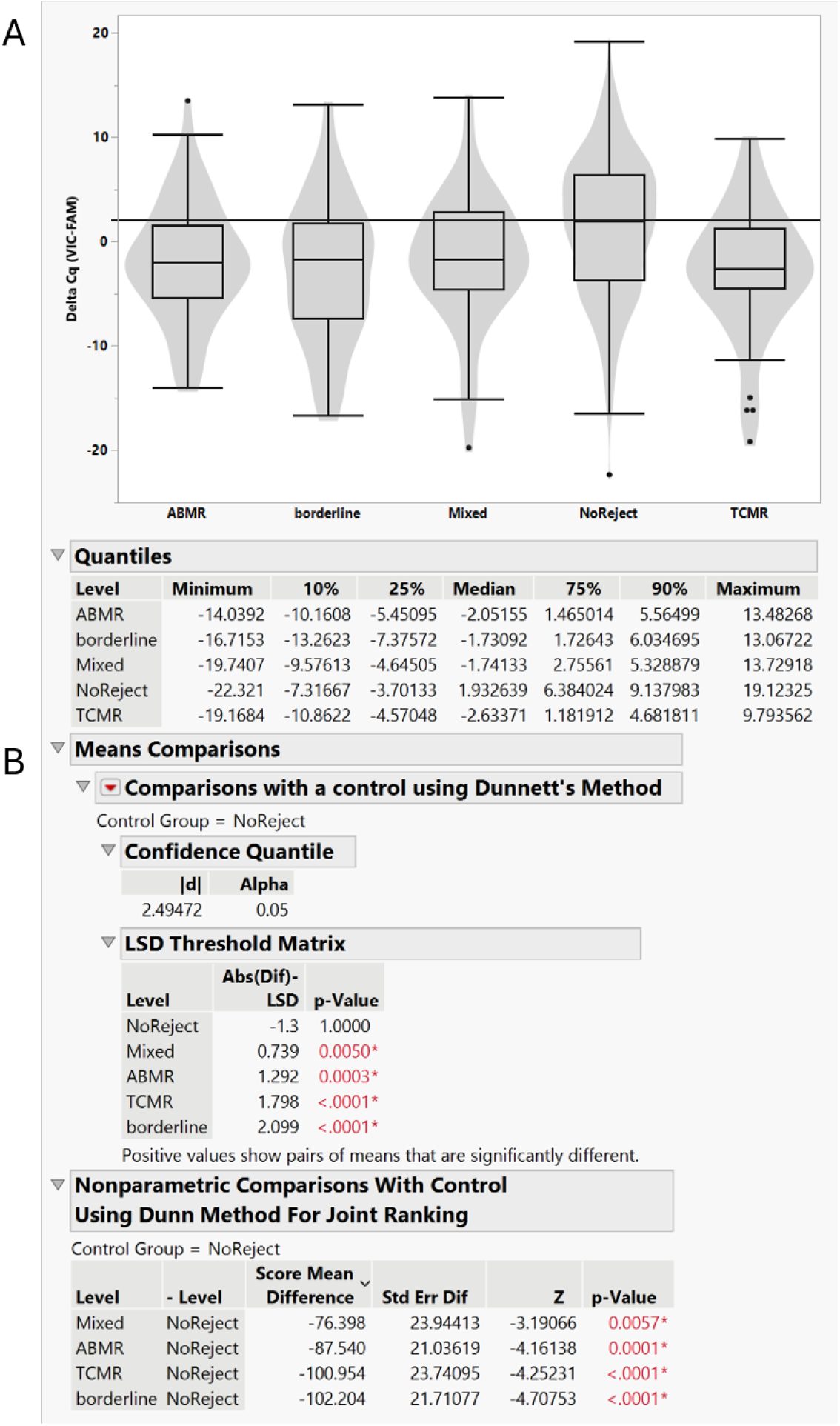
Comparing ΔCq(VIC-FAM) in different types of rejection. (A) Box plots and quantiles for each type of rejection, and (B) comparison of non-rejection group with each type of rejection by Dunnett’s method and Dunn method for joined ranking.

### 3.5 Changes in TTV/cfDNA ratios in the urine of kidney transplant patients associated with changes in biopsy results

Though significant association of TTV and cfDNA levels in urine with kidney rejection was shown in this study by several analysis methods, defining valid predictive cutoffs for ΔCq values or generating reliable risk scores based on a predictive model may require additional data for optimization and validation. Adjustment of the analytical model for the type of treatment and other clinical variables not available or not sufficiently represented in this study may also be beneficial to improve prediction. Nevertheless, qPCR technology evaluated in this study allows more frequent testing than may be feasible for more invasive or higher cost methods. To test the hypothesis that changes in TTV/cfDNA ratio in urine of kidney transplant patients may correlate with developing rejection, we analyzed qPCR data obtained from selected patients at different time post-transplant.

More than one biopsy was performed for over 25% of the patients involved in the study, and qPCR data associated with different biopsy results for each patient were analyzed in matched pairs test. Different paired results for each patient were split into 4 categories considering biopsy report and chronological order of sample collection. The first group included data for cases when the first and second biopsy did not show rejection (73 cases), the second group included pairs of samples where the first biopsy did not show rejection but the follow-up biopsy performed at a later time demonstrated active rejection (38 cases), the third group included 27 cases when rejection detected in a first biopsy was apparently successfully treated as the next biopsy performed at a later time did not show rejection, and the last group included cases not resulting in recovery from rejection since both the first and second sample were associated with biopsy indicating active rejection (67 cases). When more than 2 biopsy tests were performed for a patient, each chronologically matched pair was analyzed independently from prior or follow-up tests and only specific biopsy outcome was taken into consideration for categorization of each pair of the samples.

The delta Cq(VIC-FAM) reflecting the ratio of TTV to total cfDNA was compared in matched pairs test for each of these groups. Pairs of samples not associated with changes in patient’s condition did not show significant difference in matched pairs test with similar mean differences and confidence intervals. For 73 cases when both biopsies did not show rejection mean difference was -0.34 (CI from -1.98 to 1.31), p-value 0.68, and for 67 cases of continuous rejection mean difference was 0.19 (CI from -1.25 to 1.62), p-value 0.80. In contrast, despite the smaller sample size, groups of paired samples reflecting changes in the patient condition demonstrated significant difference in both paired t-test and non-parametric analysis (Figure S6). For 38 cases of developing rejection ΔCq(VIC-FAM) declined, with a mean difference of -3.17 (CI, from -5.55 to -0.79), p-value 0.0106 in paired t-test, and p-value 0.0143 in Wilcoxon signed rank test.

Remarkably, despite the smallest sample size, with only 27 cases of documented recovery from active rejection, this group demonstrated the most significant and consistent change in ΔCq(VIC-FAM) with a mean ΔCq increase of 4.51 (CI from 1.25 to 7.77), p-value 0.0087 in paired t-test and p-value 0.0088 in Wilcoxon signed rank test. These results indicate that changes in kidney transplant condition are associated with detectable changes in the ratio of TTV to total cfDNA in patient’s urine. It is particularly interesting that successful recovery from rejection was a change most clearly reflected in qPCR results. Detectable elevation of TTV levels in urine could be associated with increased immunosuppression resulting from treatment intervention, while reduced cfDNA in urine could reflect a reduced level of inflammation and kidney damage. Under this assumption, larger dispersion of the data observed in cases of newly developed rejection could be explained by insufficient immunosuppression levels in some patients even before rejection was detected, as a majority of the paired cases showing no changes or increase in ΔCq for the samples associated with newly developed rejection had very low levels of TTV in initial tests, when rejection was not detected by biopsy in these patients. Thus, monitoring TTV and total cfDNA in urine may reveal changes indicative of either insufficient immunosuppression or active rejection in kidney transplant patients. Further research is needed to fully understand mechanisms underlining the dynamic of TTV and cfDNA in urine of kidney transplant patients.

### 3.6 Measuring BK-virus in urine samples, combined analysis of BKV, TTV, total cfDNA and their possible correlation with biopsy results

Previous studies have demonstrated that increased levels of TTV in patient blood are associated with an increase in the risk of infections [12–14] and malignancies [15]. Similarly, Lago et al. (2024) demonstrated a significant association of TTV levels in the urine of kidney transplant patients with infections [26]. Therefore, it is important to understand how levels of TTV and cfDNA in urine may be impacted by active infection. Though clinical data associated with urine samples available for this study were limited to biopsy results, residual samples could be used to test for some infectious agents commonly detectable in the urine of transplant patients.

BK Polyomavirus (BKV) is an opportunistic viral infection, which impacts ∼30-60% of kidney transplant cases. BK polyomavirus associated nephropathy (BKPyVAN) is one of the leading causes of kidney damage in transplant patients. Currently available tests allow to detect BKV in blood, plasma or urine samples. Monitoring BKV in urine was reported as a possible method for early detection of BK-polyoma [56], but despite it’s apparent sensitivity, this method is considered insufficiently specific, and treatment changes are only recommended based on clinically validated plasma levels of BKV [57]. Availability of data for SV40 staining detecting active BKPyVAN in kidney biopsy analysis made BKV an attractive target for this follow-up study.

To evaluate whether levels of TTV in the urine of kidney transplant patients correlated with BKV and whether combined analysis of BKV, TTV and cfDNA may provide improved model of kidney rejection, residual DNA from 523 urine samples originally used for TTV and cfDNA quantification were analyzed with BKV qPCR assay (Table S6). For 21 samples, the residual amount of DNA was insufficient for additional testing, and these samples could not be included in BKV quantification experiments.

A commercially available BKV quantification qPCR test was used as recommended by the manufacturer. This method demonstrated measurement linearity within a range of concentrations spanning over eight log10 and the ability to detect single copies of DNA in qPCR reactions (Figure S7). Among 523 tested samples, 186 contained detectable levels of BKV (35.6%). Among 326 tested patients, 140 (43%) showed at least one BKV positive test, which is in agreement with clinical data previously published for kidney transplant patients, suggesting that the sample preparation and analysis process used in the study provides results comparable to common practices [58–62]. BKV quantification showed similar ratio of BKV to total cfDNA, measured as ΔCq(VIC-BKV), among all tested DNA extraction methods confirming that the method of EV concentration used for this study allows BKV extraction with efficiency comparable to commercially available kits (Figure S1).

According to the biopsy data available for the tested samples, BKPyVAN was detected in 17 non-rejection and in 5 rejection cases. A significant increase in BKV reflected by lower Cq values was observed in BKPyVAN-associated samples with median Cq 18.1 and an interquartile range from 13.6 to 22.2 while the lower quartile for BKV Cq in non-polyoma samples was 35.7. Despite a relatively small number of BKPyVAN positive samples, two tailed t-test and non-parametric Wilcoxon test demonstrated significant difference in BKV Cq between BKPyVAN positive and BKPyVAN negative groups with p-values< 0.0001. Since over 50% of non-polyoma samples did not generate Cq values in BKV qPCR, and for the purpose of statistical analysis an arbitrary value of 46 was given to all samples that did not generate Cq, results of the t-test could be impacted by this limitation. Nevertheless, excluding all BKV negative samples from data analysis did not eliminate difference observed for BKPyVAN group. Analyzing 186 BKV positive samples also demonstrated significant difference in qPCR results for BKPyVAN with both rejection and non-rejection groups (Figure S8). However, no difference between rejection and non-rejection groups was observed when Cq(BKV) or BKV qPCR results normalized by total cfDNA, ΔCq(VIC-BKV) were analyzed (Figure S8).

Previous studies evaluating correlation between levels of TTV and BKV in transplant patients did not reach a consensus: while TTV is generally considered a surrogate biomarker for the risk of diseases caused by opportunistic infections [63,64], several studies did not show significant correlation of BKV and TTV [14, 65–68]. Interestingly, Lago et al. 2024 demonstrated significant association of TTV levels between urine and plasma, however, in this study, only TTV in plasma showed significant association with the risk of viral infection but not TTV in urine [26]. Among 186 samples containing measurable levels of BKV (127 non-rejection and 59 rejection samples), qPCR results for BKV quantification did not show a significant correlation with TTV in either rejection or non-rejection groups. No significant difference between rejection and non-rejection was observed for BKV quantification (Figure S9A) with a p-value of 0.45 in two-tailed t-test. Normalizing BKV results by total cfDNA and quantifying ΔCq did not improve the association of BKV with rejection, and no significant difference between rejection and non-rejection groups was detected (p-value 0.78, Figure S9B). Nevertheless, the difference in ΔCq(VIC-FAM) reflecting TTV to cfDNA ratio was still statistically significant when rejection and non-rejection groups were compared in this subset of DNA samples (Figure S9C).

Logistic regression adjusted by patient’s sex, race, age at the time of transplant and the number of days between transplant and sample collection did not show significant association of BKV levels in urine with organ rejection (Figure S9D). In contrast, including ΔCq(VIC-FAM) to this model confirmed independent association of TTV to cfDNA ratio (Figure S10A) with the risk of rejection with p-value 0.00001 and adjusted odds ratio 0.87 per each ΔCq (CI, 0.81 – 0.93) (Figure S10B). Area under ROC curve was 0.8 (CI, 0.70 – 0.85) (Figure S10C). To evaluate whether improved odds ratio and AUC value are associated with any specific type of rejection presented in BKV positive subset of samples, logistic regression for multiple rejection types was performed (Figure S11). Interestingly, though the model evaluating total rejection events did not show significant association between rejection and time post-transplant (Figure S11A), analysis by rejection type demonstrated statistical significance for two factors, ΔCq(VIC-FAM) with p-value 0.00026 and number of days post-transplant with p-value 0.01 suggesting association between frequency of different types of rejection and time post-transplant (Figure S11A).

Predictive discrimination of resulting model was still relatively high despite the limited sample size, and AUROC was in the range from 0.79 to 0.83 for different types of rejection (Figure S11B). This level of discrimination between different types of rejection was not expected since TTV/cfDNA ratio and total cfDNA did not show significant association with specific rejection type in previous analysis. Nevertheless, adjusting the model by levels of BKV and time post-transplant could contribute to observed discrimination between different types of rejection in BKV positive samples, though the smaller sample size and relatively low event frequency could cause some inflation of AUROCs in this analysis.

Since only 35.6% of samples contained detectable levels of BKV, additional data evaluation was performed on all 523 samples using BKV measurement results as a binomial categorical variable. The BKV positive/negative category was introduced into data analysis, assigning BKV positive definition for all samples generating Cq values below 40. Categorical analysis demonstrated significant association of BKV negative category with all types of rejection with exception of TCMR. Two-sided p-value in Fisher’s exact test was 0.0027 where BKV positive group showed increase in frequency of non-rejection and TCMR and reduced frequency of all other types of rejection.

Considering the inverse association of detectable BKV with certain types of rejection, adjustment by the BKV positive/negative category was implemented in the TTV/cfDNA based neural model using 418 randomly selected samples for training (Figure 6A) and 105 for validation (Figure 6B). AUROC for both the training and validation data was over 0.8 indicating relatively good fit of a combined neural model (Figure 6). Higher AUROC values for both training and validation subsets suggested possible improvement in discrimination between rejection and non-rejection samples using model combining analysis of TTV, BKV, total cfDNA and fraction of the longer cfDNA fragments in context of time post-transplant. Demographic factors such as sex, race and age of patient at the time of transplant were not used to construct this neural model in order to reduce risk of overfitting, however, these factors may be included with availability of sufficient volume of data. Additional biomarkers, such as specific inflammatory markers in urine measured by RNA or protein quantification methods, may also find future use in combined analysis to further enhance capability of the model to discriminate between rejection and non-rejection using urine samples collected from kidney transplant patients.

**Figure 6.**
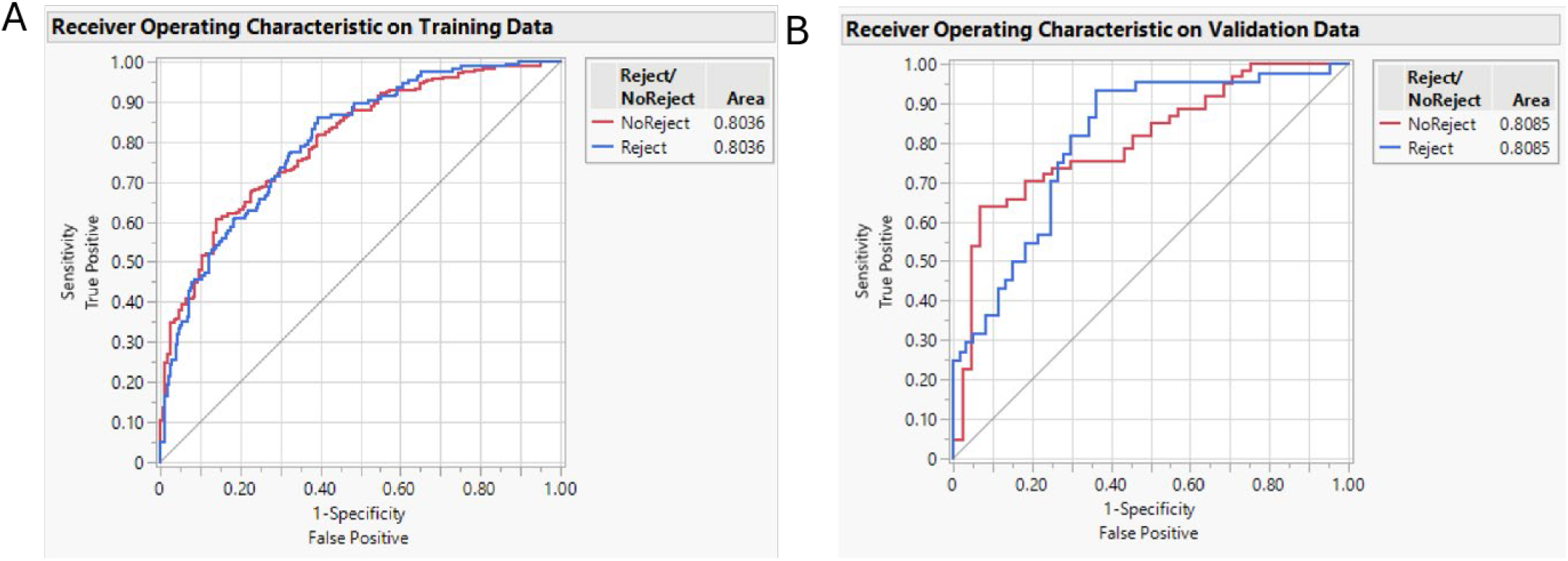
Boosted neural model for kidney rejection using TTV and cfDNA measurement data, and BKV positive/negative categorization. (A) AUROC for training and (B) AUROC for validation data sets. Model is generated by JMP Pro using qPCR results for 523 urine samples collected from 326 kidney transplant patients, in addition to qPCR data model includes number of days between transplant and sample collection. Data stratification into 418 training and 105 validation sets was performed using JMP Pro randomization algorithm.

It is interesting that information about presence or absence of BKV in urine samples can improve a predictive model for organ rejection though association of BKV levels with rejection is weak and rejection-type dependent. This result created a new question: can the absence of direct correlation between TTV and BKV levels in urine be beneficial for testing? If TTV demonstrated significant association with rejection, and BKV is an indicator of BKPyVAN, and analysis of both these markers can be further enhanced by measuring cfDNA in urine samples, would the combination of these biomarkers improve discrimination between rejection and BKPyVAN?

Previous studies demonstrated that increase of BKV in urine can detect BKPyVAN development earlier than blood tests [56], however, relatively low positive predictive value for urine test presents a risk of false positive BKPyVAN results in rejection cases [57,62,69]. Since BKPyVAN is commonly treated by reducing immunosuppression, it is particularly important to differentiate BKPyVAN and rejection. Combined analysis of qPCR results for TTV, cfDNA and BKV was performed to assess feasibility to improve predictive values in differentiating BKPyVAN and rejection from normal samples.

Initial data analysis performed in this study defined samples based on presence or absence of biopsy-proven rejection. All BKPyVAN related samples were allocated to rejection (5 samples) or non-rejection category (17 samples) in initial analysis. To evaluate feasibility of differentiating rejection from BKPyVAN, samples were separated into 3 groups, rejection, BKPyVAN and non-rejection. Biopsy results for five samples collected from 4 patients demonstrated combination of BKPyVAN and active rejection. Since group representing this mixed condition was too small, samples were excluded from following analysis. Only non-rejection BKPyVAN cases were analyzed as described below.

To further evaluate association of different variables with each of three outcomes and assess feasibility to develop a model capable of discriminating BKPyVAN and rejection using urine biomarkers, several nominal logistic regression models were created. All models were adjusted by age of the patient at time of transplant and number of days between transplant and sample collection. In addition to age and time interval, model 1 included Cq(BKV); model 2 included ΔCq(VIC-BKV) for BKV/cfDNA ratio analysis; model 3 used ΔCq(VIC-FAM) for TTV/cfDNA ratio analysis; model 4 included ΔCq(VIC-FAM), total cfDNA, fraction of longer DNA fragments and BKV positive/negative categorization; and model 5 included full combination of all qPCR results for TTV, cfDNA and BKV quantification data. ROC and PRC curves for all 3 tested outcomes were evaluated (Figure 7).

**Figure 7.**
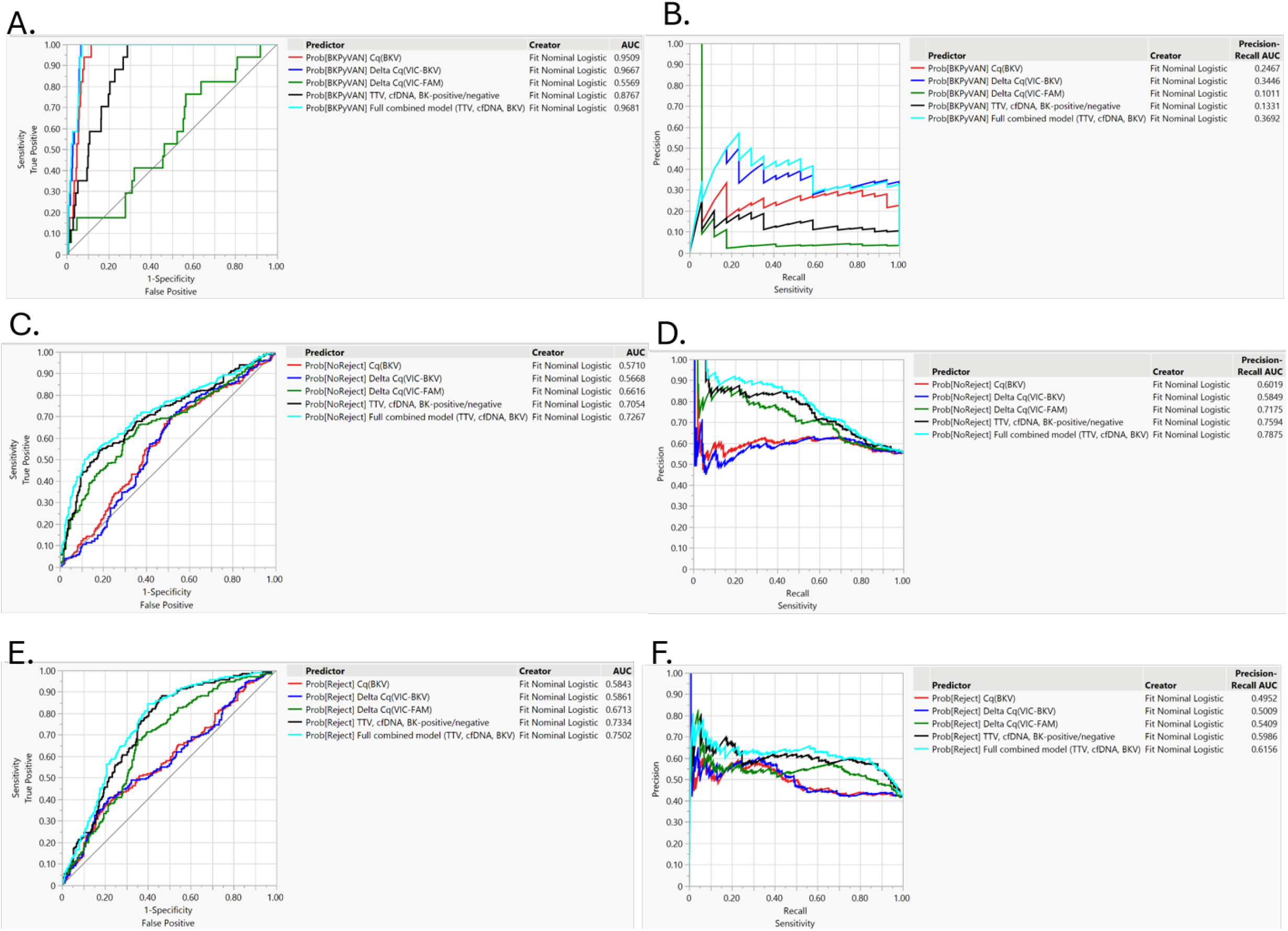
Comparing five nominal logistic models evaluating different combinations of covariates. (A) ROC curves and AUROC values for BKPyVAN, (B) PRC curves and AUPRC values for BKPyVAN, (C) ROC curves and AUROC values for normal outcome (NoReject), (D) PRC curves and AUPRC values for normal outcome (NoReject), (E) ROC curves and AUROC values for rejection, (F) PRC curves and AUPRC values for rejection. All models were adjusted by patient’s age at the time of transplant and number of days between transplant and sample collection.

Nominal logistic regression analyzing association of Cq(BKV) with BKPyVAN and rejection was compared with model using ΔCq(VIC-BKV). Models were adjusted by age of the patient at the time of transplant and the number of days between transplant and sample collection. Analysis demonstrated clear discrimination of BKPyVAN from either rejection or non-rejection samples with AUROC over 0.9 for both models. DeLong AUROC comparison method [40] showed that model using ΔCq(VIC-BKV) provided stronger predictive discrimination in comparison to model generated with Cq(BKV) not adjusted by cfDNA concentration (Figure S12).

Using ROC curves, which reflect only sensitivity and specificity, for model characterization is considered more appropriate for well-balanced data sets, because AUROC can give an overly optimistic impression of clinical utility when the positive outcome is uncommon in the investigated population [70]. Since the number of polyoma related samples was relatively low (3.3% of total samples), ROC curves may be insufficient to fully characterize each relevant biomarker within this study. For this reason, not only ROC but also precision recall curves (PRC) were assessed for resulting models. Comparing model 1 and model 2 showed highly similar AUROC values for BKPyVAN outcome: 0.95 for Cq(BKV) and 0.97 for ΔCq(VIC-BKV), nevertheless, the difference in PRC shapes was much more noticeable (Figure S12). Area under precision recall curve (AUPRC) for Cq(BKV) model was 0.25 (CI, 0.14 – 0.42) and for ΔCq(VIC-BKV) model AUPRC was 0.34 (CI, 0.19 – 0.59) for BKPyVAN confirming improvement of predictive discrimination for BKV/cfDNA ratio. As expected, BKV quantification results did not have significant association with rejection and no significant difference between Cq(BKV) and ΔCq(VIC-BKV) in AUROC for rejection was observed.

Five nominal logistic models including different combinations of candidate predictors were evaluated for all three outcomes. Though rejection and non-rejection related samples are relatively well balanced in analyzed cohort, AUPRC values were reported for each model and each outcome (Table 3). Analysis showed that a model combining all qPCR data for TTV, BKV and cfDNA demonstrated highest AUC values for both ROC and PRC curves in all 3 outcomes (Table 3). All 5 models were analyzed using DeLong AUROC comparison method [40] (Table 4). As expected, models including BKV data showed discrimination for BKPyVAN, and models including TTV and cfDNA showed improved discrimination of rejection outcome. Nevertheless, an additional increase in AUC for rejection was also observed as a contribution of BKV data. Improved discrimination was particularly apparent for the rejection group, where replacing the BKV positive/negative categorization by ΔCq(VIC-BKV) led to significantly increased AUROC when model 4 and model 5 were compared, as evidenced by non-parametric comparison (p-value 0.0119) (Table 4). Difference between full quantitative model and ΔCq(VIC-FAM) was even more apparent (p<0.0001 for rejection) indicating significant contribution of total cfDNA and fraction of longer fragments in addition to BKV quantification for discriminating rejection samples from two other outcomes.

**Table 3.** Model quality comparison of 5 nominal logistic models analyzing three possible outcomes: normal biopsy, BKPyVAN and rejection.

| Outcome<br>(% of total) | Model analyzed<sup>a</sup> | AUROC Analysis |  |  | AUPRC Analysis |  |  |
| --- | --- | --- | --- | --- | --- | --- | --- |
|  |  | AUC<sup>b</sup> | Lower<br>95% | Upper<br>95% | AUC<sup>c</sup> | Lower<br>95% | Upper<br>95% |
| BKPyVAN (3.3) | Cq(BKV) | 0.95 | 0.93 | 0.97 | 0.25 | 0.14 | 0.42 |
| BKPyVAN (3.3) | ΔCq(VIC-BKV) | 0.97 | 0.95 | 0.98 | 0.34 | 0.19 | 0.59 |
| BKPyVAN (3.3) | ΔCq(VIC-FAM) | 0.56 | 0.42 | 0.68 | 0.10 | 0.02 | 0.25 |
| BKPyVAN (3.3) | TTV, cfDNA, BK-positive/negative | 0.88 | 0.82 | 0.92 | 0.13 | 0.07 | 0.26 |
| BKPyVAN (3.3) | Full quantitative model (TTV, cfDNA, BKV) | 0.97 | 0.95 | 0.98 | 0.37 | 0.20 | 0.63 |
| NoReject (55.4) | Cq(BKV) | 0.57 | 0.52 | 0.62 | 0.60 | 0.54 | 0.66 |
| NoReject (55.4) | ΔCq(VIC-BKV) | 0.57 | 0.52 | 0.62 | 0.58 | 0.53 | 0.65 |
| NoReject (55.4) | ΔCq(VIC-FAM) | 0.66 | 0.61 | 0.71 | 0.72 | 0.66 | 0.77 |
| NoReject (55.4) | TTV, cfDNA, BK-positive/negative | 0.71 | 0.66 | 0.75 | 0.76 | 0.71 | 0.81 |
| NoReject (55.4) | Full quantitative model (TTV, cfDNA, BKV) | 0.73 | 0.68 | 0.77 | 0.79 | 0.74 | 0.83 |
| Reject (41.3) | Cq(BKV) | 0.58 | 0.53 | 0.63 | 0.49 | 0.43 | 0.56 |
| Reject (41.3) | ΔCq(VIC-BKV) | 0.59 | 0.54 | 0.64 | 0.50 | 0.43 | 0.57 |
| Reject (41.3) | ΔCq(VIC-FAM) | 0.67 | 0.62 | 0.72 | 0.54 | 0.47 | 0.61 |
| Reject (41.3) | TTV, cfDNA, BK-positive/negative | 0.73 | 0.69 | 0.77 | 0.60 | 0.53 | 0.67 |
| Reject (41.3) | Full quantitative model (TTV, cfDNA, BKV) | 0.75 | 0.71 | 0.79 | 0.62 | 0.55 | 0.69 |

<sup>a</sup> All models included patient's age at the time of transplant and number of days between transplant and sample collection in addition to parameters listed in the table
<sup>b</sup> AUROC and confidence intervals were calculated by JMP Pro using DeLong method
<sup>c</sup> AUPRC were calculated in JMP Pro using standard model analysis process, and 95% confidence intervals were calculated by JMP using bootstrap method with 2500 sampling runs.

**Table 4.**
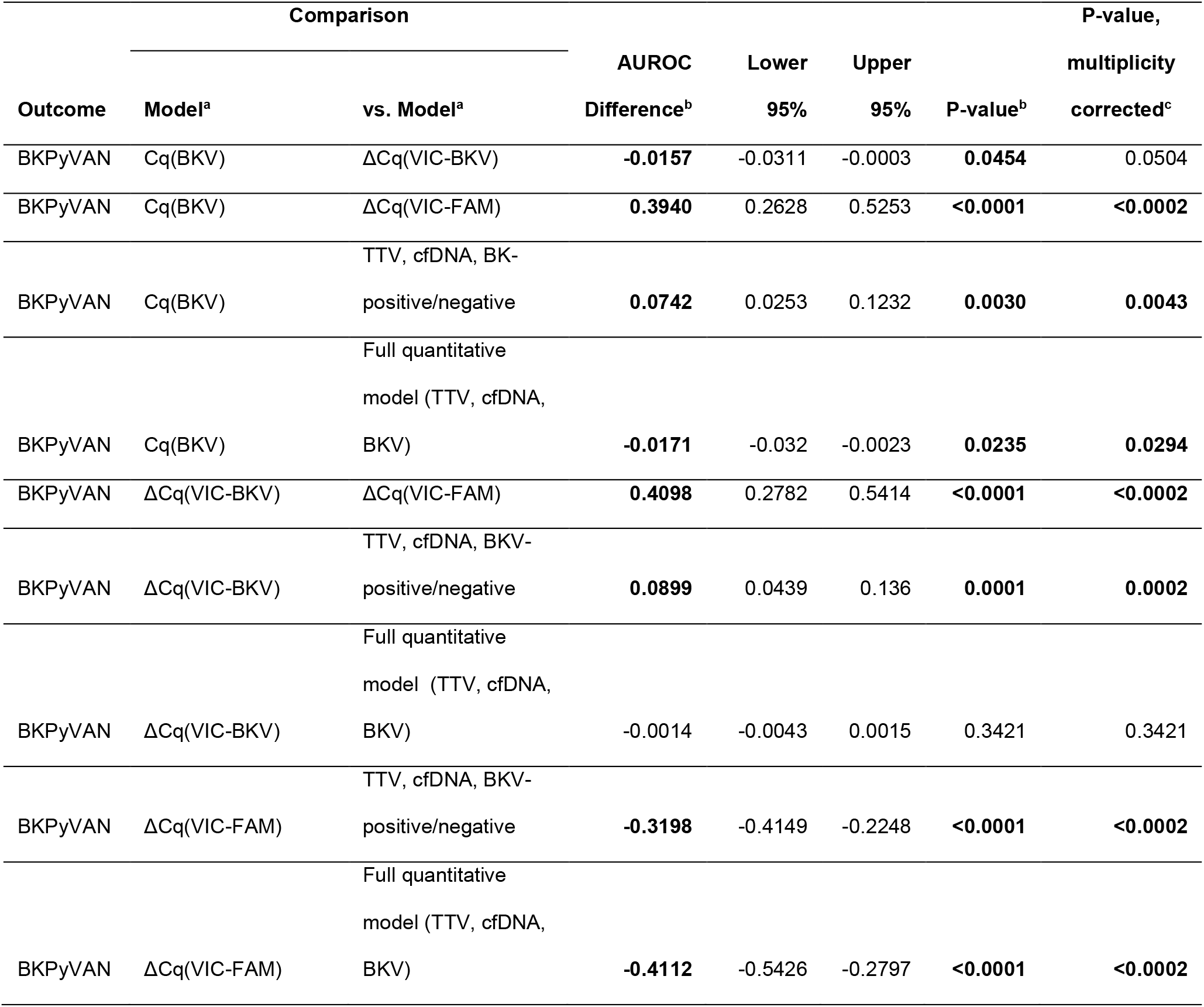
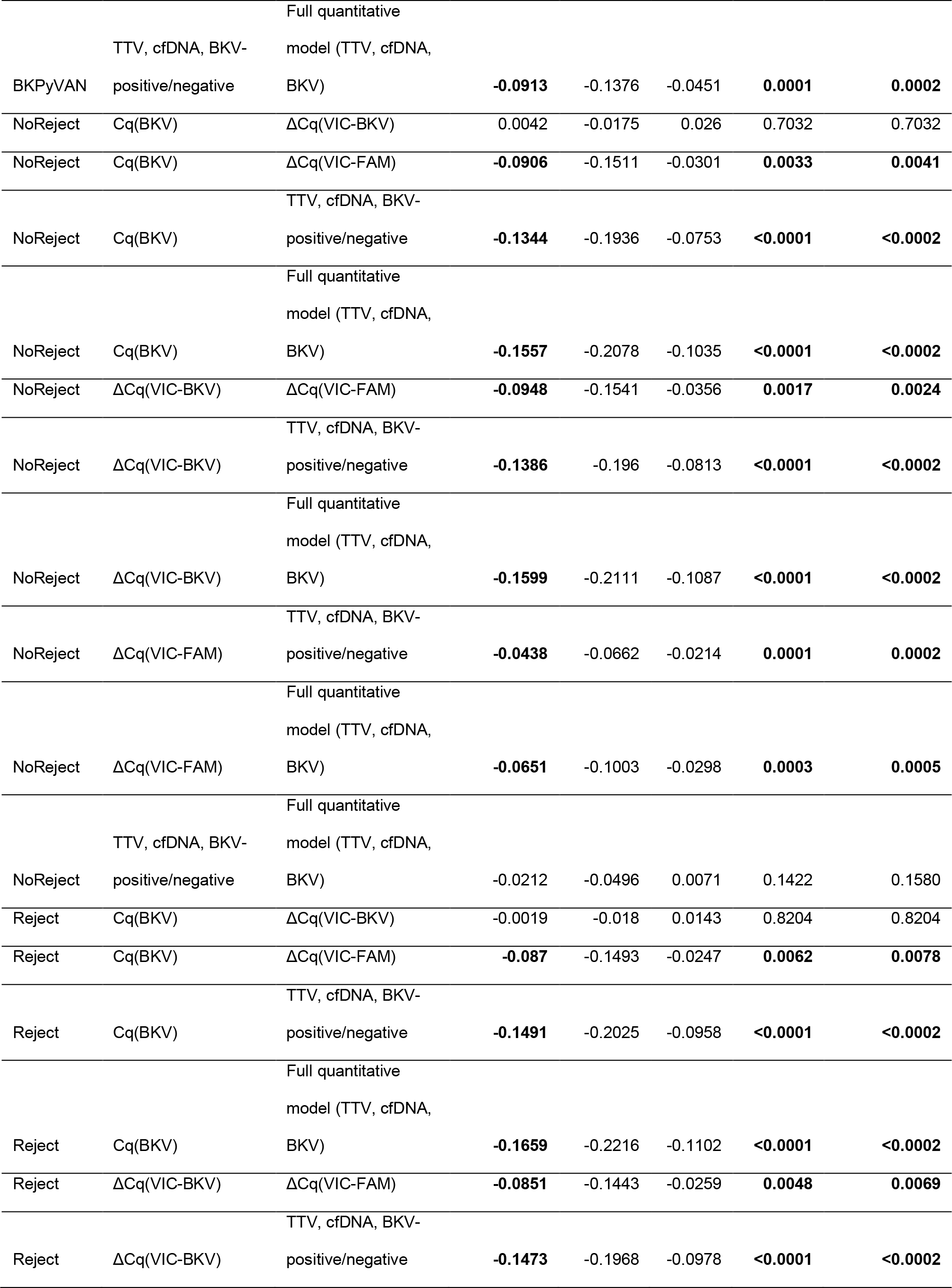
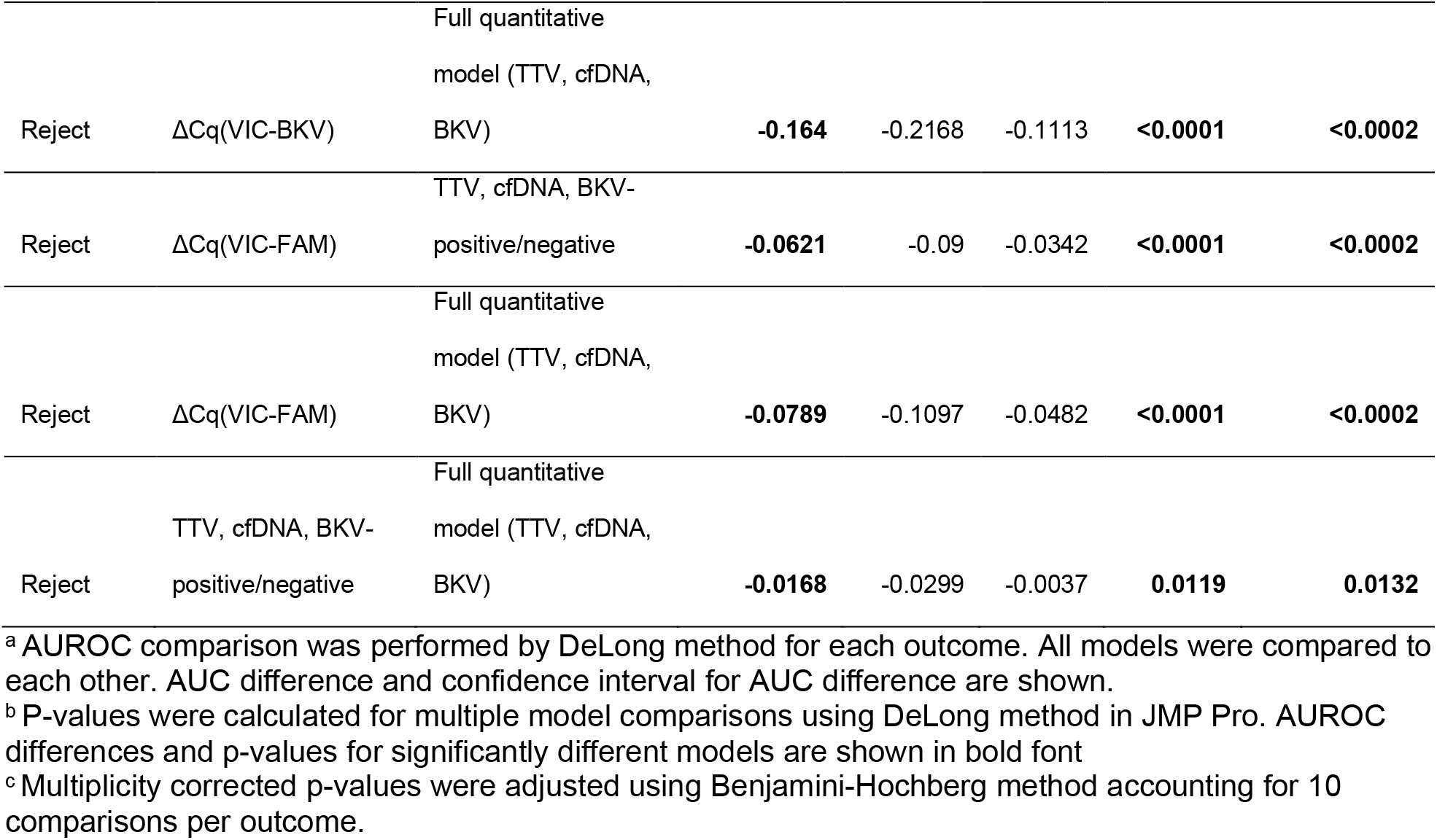
Model comparison analysis.

To reduce the risk of an analysis error in multiple comparisons, p-values were evaluated after adjustment by the Benjamini-Hochberg method (Table 4). The impact of this adjustment was noticeable only for the BKPyVAN outcome, likely due to the low event frequency. Interestingly, though AUROC difference for BKPyVAN between Cq(BKV) and ΔCq(VIC-BKV) became insignificant after p-value adjustment for multiple comparisons, difference between Cq(BKV) and full quantitative model remained significant even after Benjamini-Hochberg correction (Table 4). Data analysis supported the hypothesis that combining TTV, BKV, cfDNA and level of DNA fragmentation into a single model may provide the ability to discriminate different biopsy-confirmed outcomes by a urine test. Due to a small number of samples associated with confirmed BKPyVAN we did not attempt advanced modelling for 3-outcome classification, however continuing investigation of urine biomarkers can further improve methods of post-transplant monitoring, differentiating infections from rejection in kidney transplant patients. Results of this study were completely aligned with initial expectations. Lack of correlation between TTV and BKV suggested that feasibility of using TTV as a surrogate marker for the risk of opportunistic infections may not be applicable to some viruses or some types of samples. Results of this study also suggest that changes in TTV levels associated with organ rejection may be different among biological samples due to possible impact of local inflammatory processes and lymphocyte infiltration. Significant inverse association of TTV/cfDNA ratios with kidney rejection was attributed to concurrent decrease in TTV and increase in total cfDNA in urine resulting from inflammatory processes, however, this study could not assess whether the observed phenomenon was unique for urine or could also be detectable in blood.

The main limitations of this study include insufficient information regarding patient treatment and treatment adjustments over duration of time. Infectious diseases information is also limited by biopsy confirmed BKPyVAN, no other information regarding opportunistic infections or developing malignancies is available for the patients participating in this study limiting feasibility to correlate TTV in urine with overimmunosuppression. We were able to test our hypotheses only partially, showing that TTV and cfDNA levels in urine have significant association with rejection, however, we were not able to compare levels of each biomarker in blood to urine. This comparison could not be carried out because no matching blood samples were available, and we still cannot conclude whether active rejection causes local reduction of TTV levels in the impacted organ or levels of TTV are generally reduced and can be monitored in blood with comparable correlation to the outcomes. An additional limitation of the study is the possible presence of cellular debris contaminating the EV fraction, since urine samples were initially frozen without centrifugation, and some cellular particles could be released from cells during freeze/thaw prior to urine pellet removal.

TTV and cfDNA analysis used in this study was performed by an in-house qPCR assay intended for research use only and not approved for clinical use by FDA or any other organization. This test was not commercially available at the time of the study. Additional studies are needed to further evaluate the associations of urinary TTV, BKV, and cfDNA with kidney allograft rejection and BKPyVAN.

## Data Availability

All data produced in the present study are available upon reasonable request to the authors after patent submission process is completed

## Acknowledgments

We would like to express our gratitude to Dr. Nickolai Alexandrov for valuable discussion of statistical analysis and predictive modeling approaches.

## Funding

This study was fully funded by Thermo Fisher Scientific Inc.

## Informed Consent Statement

Additional informed consent is not required for this study as it was obtained by the sample provider, Verici Dx plc, who followed IRB protocols with informed consent, and provided de-identified samples. Samples were collected by Verici Dx under VDX 20-1001 study, which was approved by the Advarra Institutional Review Board (Pro00049177), all participants provided written informed consent. Participants were enrolled in this study from March 2021 to January 2023.

## Institutional Review Board Statement

VDX 20-1001 study was approved by the Advarra Institutional Review Board (Pro00049177), and all participants provided written informed consent. Participants were enrolled in this study from March 2021 to January 2023. Samples used in this study were deidentified. Investigators did not contact the subjects and did not re-identify or attempt to re-identify them.

## Data Availability Statement

The data discussed in this study are available on request from the corresponding author upon completion of an ongoing patent application.

## Author contributions

Conceptualization, A.K.C.; methodology, Y.A., Z.W., M.M., J.C., A.C. and A.K.C.; data acquisition, Y.A., Z.W., M.M., J.C., A.C; experimental analysis, Y.A., Z.W., M.M., and A.K.C.; formal analysis, Y.A, and A.K.C.; investigation, Y.A., M.M., Z.W., J.C., A.C. and A.K.C.; resources, A.V. and A.K.C.; data curation, Y.A., M.M., Z.W., and A.K.C.; writing—original draft preparation, A.K.C.; writing—review and editing, Y.A., Z.W., A.C., A.V., J.C. and A.K.C. All authors have read and agreed on the contents of the manuscript.

## Consent for publication

Not applicable.

## Conflict of interest

Authors are current or former employees of One Lambda Inc., part of Thermo Fisher Scientific Inc., which submitted a patent application covering concepts disclosed in this manuscript. The submitted application is not published as of the date of this submission.

## Declaration of generative AI and AI-assisted technologies in the writing process

During the preparation of this work, the authors used a company-licensed version of OpenAI’s GPT-4 model, integrated within the Gene.AI platform provided by Thermo Fisher Scientific, in order to ensure that paragraphs were free of grammatical errors and to improve readability. After using this tool, the authors reviewed and edited the content as needed and take full responsibility for the content of the publication.

## Notes

### Competing Interest Statement

The authors have declared no competing interest.

### Author Declarations

Samples collected by Verici Dx were part of the VDX 20-1001 study, which was approved by the Advarra Institutional Review Board (Pro00049177), and all participants provided written informed consent.

