## Supplemental figures and legends for "To TTV or Not to TTV? A Combined Urinary Biomarker Approach in Kidney Transplant Recipients Using Torque Teno Virus, Cell-Free DNA, and BK Polyomavirus Measurements": Supplemental figure legends.pdf

**Figure S1.** Comparison of methods for cfDNA extraction from urine. Three DNA extraction methods were compared for their ability to extract cfDNA, TTV and BKV DNA from human urine. A first method, named EV+viral, is an in-house sample preparation method comprising concentration of extracellular vesicles by precipitation followed by viral sample preparation using the EZ1&2 Virus Mini kit v2.0. The in-house method was tested on 6 randomly selected human samples side by side with cfDNA extraction methods by Norgen and Qiagen following manufacturer's recommendations. For all 3 methods, 1 mL of urine was used. (A) qPCR results for TTV, SHORT and BGIC markers were analyzed, ΔCq was for 6 human samples to assess ratio of TTV to cfDNA (ΔCq(VIC-FAM)) and the fraction of longer cfDNA fragments (ΔCq(VIC-JUN)). (B) One-way analysis of ΔCq(VIC-FAM) in samples extracted by 3 different methods using t-test and non-parametric tests comparing different methods. (C) Comparison of BKV extraction results for three different sample preparation methods, Cq(BKV) and ratio of BKV/cfDNA as delta Cq(VIC-BKV) are analyzed.

**Figure S2**. Analysis of data from the confirmatory study of 300 urine samples (107 rejection and 193 non-rejection). (A) Comparison of ΔCq(VIC-FAM) in rejection and non-rejection samples using two-tailed t-test and non-parametric test. (B-C) Nominal logistic regression analysis of the association between biopsy outcomes and qPCR data. Analysis is adjusted by sex, race, age at the time of transplant and time between transplant and sample collection. (B) Effect summary of the logistic model and (C) per-unit odds ratios for rejection/no-rejection odds estimated within the nominal logistic model.

**Figure S3.** Analysis of dPCR data using logistic regression. (A) Model 1 – a nominal logistic regression using log10(TTV/cfDNA), log10(BGIC/SHORT), log10(total cfDNA) and adjusted by patient sex, race, age at the time of transplant and the number of days between transplant and sample collection; (B) Model 2 - nominal logistic regression using log10(TTV/cfDNA), log10(BGIC/SHORT), log10(total cfDNA) without additional adjustments; (C) Model 3 - nominal logistic fit using only log10(TTV/cfDNA) ratio. (D) ROC graphs are shown for all 3 models. (E-F) Model comparison was performed by the DeLong method using JMP Pro software, (E) AUROC, 95% confidence intervals are shown for all three models, and (F) 95% confidence interval for AUROC differences and p-values are shown in a table.

**Figure S4.** Analysis of TTV/cfDNA ratios measured by dPCR in a confirmatory study. The log10(TTV/cfDNA) values were compared between rejection and no-rejection groups. Two-tailed t-test and non-parametric test results are shown.

**Figure S5.** Comparison of ΔCq(VIC-FAM) between samples associated with non-rejection and various types of rejection reported in biopsy results

**Figure S6.** Matched pairs analysis comparing ΔCq(VIC-FAM) in consecutive tests available for individual patients. Results of the paired t-test and non-parametric Wilcoxon signed rank test are shown. (A) Paired analysis where first and second test did not have signs of rejection. (B) Matched pair analysis where first and second test were associated with biopsy-confirmed rejection, (C) Matched pair analysis where first test was not associated with documented rejection however follow up biopsy demonstrated active rejection. (D) Matched pair analysis where first tested sample was associated with biopsy-confirmed rejection, and follow up biopsy did not show rejection for the same patient. The difference between the ΔCq(VIC-FAM) values for paired samples (Y axis) was plotted against their mean (X axis) on all presented graphs.

**Figure S7.** Standard curve and linearity analysis for the BKV quantification qPCR assay used in the study. Three replicate-reactions were tested per concentration with the exception of reactions with expected input below 100 copies per reaction, for which 10 replicates were tested. $R^2$ for linear fit of presented data and regression formula are shown. (A) Standard curve for BKV copy number estimation. BKV quantification standard was diluted from 36, 000 copies per reaction to single copies; log10 of copy number is shown on X axis and qPCR results are shown on Y axis. (B) For DNA input below 100 copies per reaction, 10 replicates were tested, Poisson distribution analysis was performed with estimates and confidence intervals for Cq(BKV) calculated, results of this analysis are shown in a table. (C) Standard curve for BKV copy number estimation with Cq(BKV) for low DNA input values are replaced by mean value for each DNA input estimated by Poisson distribution analysis. This standard curve was used to evaluate linearity range limitation while addressing sampling variability expected for low DNA input. Cq values for each replicate were used for DNA input over 20 copies per reaction, and for DNA input below 20 copies per reaction, Poisson distribution estimates for Cq(BKV) were used as a single point per concentration. The resulting formula was used to calculate log10(BKV copy/reaction) of tested human samples. (D) Linearity analysis was performed by testing series of 4-fold dilutions of DNA extracted from human urine to confirm linearity of the assay in human samples. A sample was selected for a high starting level of BKV (over 8 log10), the X axis shows log10 of sample dilutions, and the Y axis displays log10 of BKV copy number recalculated from Cq values using results from standard curve.

**Figure S8.** BKV quantification in BKPyVAN, rejection and normal urine samples, data for 186 BKV-positive samples were analyzed. (A) Box plots and contour of data distribution for ΔCq(VIC-BKV) and non-normalized Cq(BKV) associated with each outcome. (B) Comparison of ΔCq(VIC-BKV) between BKPyVAN and other samples using Dunnett’s method and (C) Comparison of Cq(BKV) between BKPyVAN and other samples using Dunnett’s method.

**Figure S9.** Analysis of BKV, cfDNA and TTV data in the BKV positive subset of samples, data for 186 BKV-positive samples were analyzed. Rejection and non-rejection groups are compared. Boxplots and two-sided p-values for t-test are shown for (A) Cq(BKV), (B) ΔCq(VIC-BKV) and (C) ΔCq(VIC-FAM). (D) Nominal logistic regression analysis adjusted by sex, race, age of the patient at the time of transplant and number of days between transplantation and sample collection was performed for Cq(BKV) to further evaluate possible association with rejection in adjusted model.

**Figure S10.** Combined logistic regression model for BKV positive samples. The model analyzed the association of candidate predictors with rejection and included all qPCR data for TTV, cfDNA and BKV. The model was adjusted by sex, race, age of the patient at the time of transplantation and number of days post-transplant. (A) Effect summary of tested covariates, (B) adjusted per unit odds ratios, (C) ROC graph with AUROC value is shown.

**Figure S11.** Combined logistic regression model for BKV positive samples. The model analyzed the association of candidate predictors with different types of rejection and included all qPCR data for TTV, cfDNA and BKV. The model was adjusted by sex, race, age of the patient at the time of transplantation and number of days post-transplant. (A) Effect summary of tested covariates, (B) ROC graphs with AUROC values are shown.

**Figure S12.** Comparison of the nominal logistic models analyzing the association of Cq(BKV) or ΔCq(VIC-BKV) with biopsy outcomes. Both models were adjusted by age of the patient at the time of transplant and the number of days between transplant and sample collection. (A) ROC graphs with AUROC values for BKPyVAN outcome, (B) AUC comparison analysis by DeLong method, (C) PRC graphs with AUPRC for BKPyVAN outcome, (D) ROC graphs with AUROC values for rejection outcome.
