## Supplemental figures and legends for "To TTV or Not to TTV? A Combined Urinary Biomarker Approach in Kidney Transplant Recipients Using Torque Teno Virus, Cell-Free DNA, and BK Polyomavirus Measurements": Supplemental figures.pdf

### Figure S1. Comparison of different DNA extraction methods

A

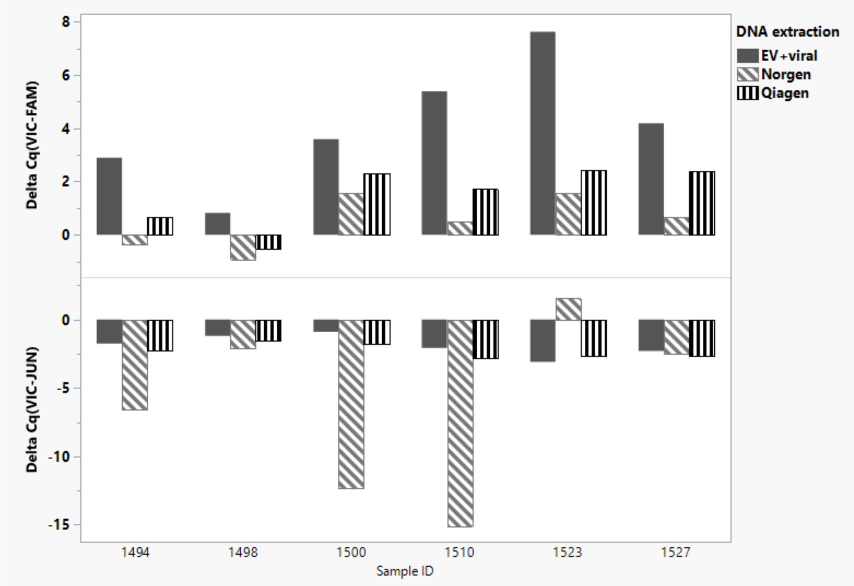


B

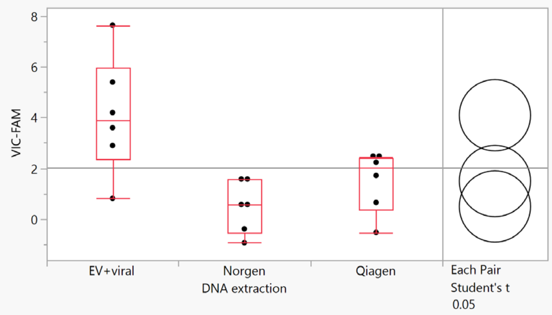


**Quantiles**

| Level | Minimum | 10% | 25% | Median | 75% | 90% | Maximum |
|---|---|---|---|---|---|---|---|
| EV+viral | 0.816619 | 0.816619 | 2.373072 | 3.890568 | 5.946474 | 7.626432 | 7.626432 |
| Norgen | -0.9271 | -0.9271 | -0.51792 | 0.575434 | 1.574326 | 1.579168 | 1.579168 |
| Qiagen | -0.51876 | -0.51876 | 0.36268 | 2.018405 | 2.389521 | 2.435297 | 2.435297 |

**Nonparametric Comparisons For Each Pair Using Wilcoxon Method**

| q* | Alpha |
|---|---|
| 1.95996 | 0.05 |

| Level | - Level | Score Mean Difference | Std Err Dif | Z | p-Value | Hodges-Lehmann | Lower CL | Upper CL | Difference Plot |
|---|---|---|---|---|---|---|---|---|---|
| Qiagen | Norgen | 3.16667 | 2.081666 | 1.52122 | 0.1282 | 0.95030 | -0.92267 | 2.75579 | |
| Qiagen | EV+viral | -4.50000 | 2.081666 | -2.16173 | 0.0306* | -2.35388 | -5.30610 | -0.16013 | |
| Norgen | EV+viral | -5.16667 | 2.081666 | -2.48199 | 0.0131* | -3.61342 | -6.31359 | -1.19815 | |

C

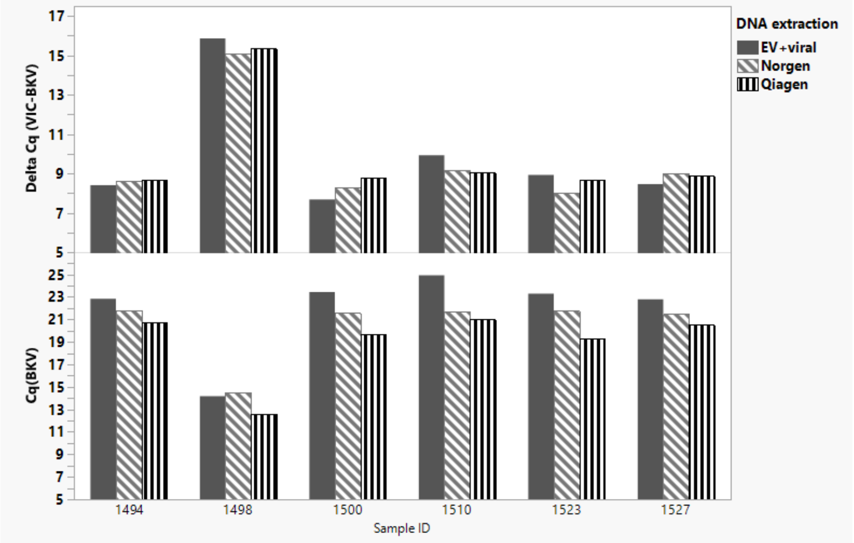

### Figure S2. Delta Cq(VIC-FAM) analysis in confirmatory data set.

A

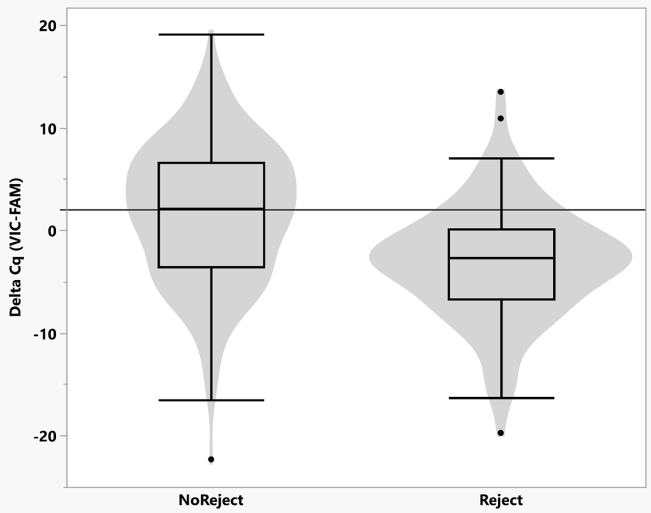


##### t Test

Reject-NoReject
Assuming unequal variances

| | | | |
|---|---|---|---|
| Difference | -4.8082 | t Ratio | -6.58957 |
| Std Err Dif | 0.7297 | DF | 258.3661 |
| Upper CL Dif | -3.3713 | Prob > \|t\| | <.0001* |
| Lower CL Dif | -6.2450 | Prob > t | 1.0000 |
| Confidence | 0.95 | Prob < t | <.0001* |

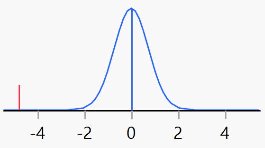


##### Wilcoxon / Kruskal-Wallis Tests (Rank Sums)

| Level | Count | Score Sum | Expected Score | Score Mean | (Mean-Mean0)/Std0 |
|---|---|---|---|---|---|
| NoReject | 193 | 33466.0 | 29046.5 | 173.399 | 6.140 |
| Reject | 107 | 11684.0 | 16103.5 | 109.196 | -6.140 |

###### Wilcoxon Two-Sample Test, Normal Approximation

| S | Z | Prob>Z | Prob>\|Z\| |
|---|---|---|---|
| 11684 | -6.13989 | <.0001* | <.0001* |

###### Kruskal-Wallis Test, ChiSquare Approximation

| ChiSquare | DF | Prob>ChiSq |
|---|---|---|
| 37.7068 | 1 | <.0001* |

B

| Source | Logworth | PValue |
|---|---|---|
| Delta Cq (VIC-FAM) | 8.312 | 0.00000 |
| Delta Cq(VIC-JUN) | 0.667 | 0.21533 |
| SEX | 0.303 | 0.49798 |
| RACE | 0.297 | 0.50461 |
| Log10[Days post-transplant] | 0.153 | 0.70386 |
| Age at Time of Transplant | 0.062 | 0.86604 |

C

| Term | Odds Ratio | Lower 95% | Upper 95% | Reciprocal |
|---|---|---|---|---|
| Delta Cq (VIC-FAM) | 0.88289 | 0.842165 | 0.922557 | 1.1326443 |
| Delta Cq(VIC-JUN) | 0.937634 | 0.849206 | 1.039476 | 1.0665142 |
| Age at Time of Transplant | 1.001579 | 0.983417 | 1.020282 | 0.9984231 |
| Log10[Days post-transplant] | 0.874293 | 0.434512 | 1.745898 | 1.143781 |

### Figure S3. Results of dPCR, comparison of nominal logistic models

A.

| Source | Logworth | PValue |
|---|---|---|
| Log10(TTV/cfDNA total) | 8.097 | 0.00000 |
| Log10(cfDNA total) | 2.023 | 0.00948 |
| Log10(BGIC/SHORT) | 1.305 | 0.04959 |
| Age at Time of Transplant | 0.468 | 0.34031 |
| Days post transplant | 0.190 | 0.64583 |
| SEX | 0.169 | 0.67708 |
| RACE | 0.154 | 0.70094 |

B.

| Source | Logworth | PValue |
|---|---|---|
| Log10(TTV/cfDNA total) | 8.684 | 0.00000 |
| Log10(cfDNA total) | 2.233 | 0.00585 |
| Log10(BGIC/SHORT) | 1.320 | 0.04783 |

C.

**Parameter Estimates**

| Term | Estimate | Std Error | ChiSquare | Prob>ChiSq |
|---|---|---|---|---|
| Intercept | -0.7569592 | 0.1547607 | 23.92 | <.0001* |
| Log10(TTV/cfDNA total) | -0.509218 | 0.1002327 | 25.81 | <.0001* |

For log odds of Reject/NoReject

D.

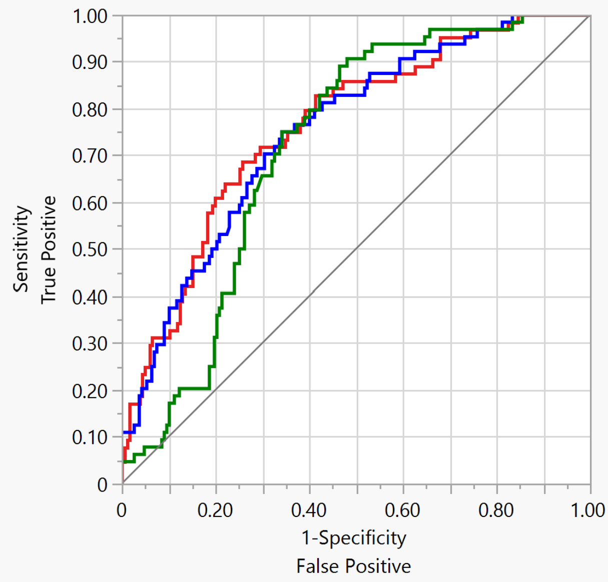


| Predictor | Creator | AUC |
|---|---|---|
| Prob[Reject] Model 1 | Fit Nominal Logistic | 0.7632 |
| Prob[Reject] Model 2 | Fit Nominal Logistic | 0.7515 |
| Prob[Reject] Model 3 | Fit Nominal Logistic | 0.7180 |

E.

**AUC Comparison for Reject/NoReject=Reject**

| Predictor | AUC | Std Error | Lower 95% | Upper 95% |
|---|---|---|---|---|
| Prob[Reject] Model 1 | 0.7632 | 0.0338 | 0.6909 | 0.8229 |
| Prob[Reject] Model 2 | 0.7515 | 0.0339 | 0.6795 | 0.8119 |
| Prob[Reject] Model 3 | 0.7180 | 0.0328 | 0.6495 | 0.7777 |

F.

| Predictor | vs. Predictor | AUC Difference | Std Error | Lower 95% | Upper 95% | ChiSquare | Prob>ChiSq |
|---|---|---|---|---|---|---|---|
| Prob[Reject] Model 1 | Prob[Reject] Model 2 | 0.0117 | 0.0119 | -0.012 | 0.0349 | 0.9656 | 0.3258 |
| Prob[Reject] Model 1 | Prob[Reject] Model 3 | 0.0452 | 0.0235 | -0.001 | 0.0912 | 3.715 | 0.0539 |
| Prob[Reject] Model 2 | Prob[Reject] Model 3 | 0.0335 | 0.0216 | -0.009 | 0.0758 | 2.4186 | 0.1199 |

### Figure S4. Results of dPCR analysis log10(TTV/cfDNA)

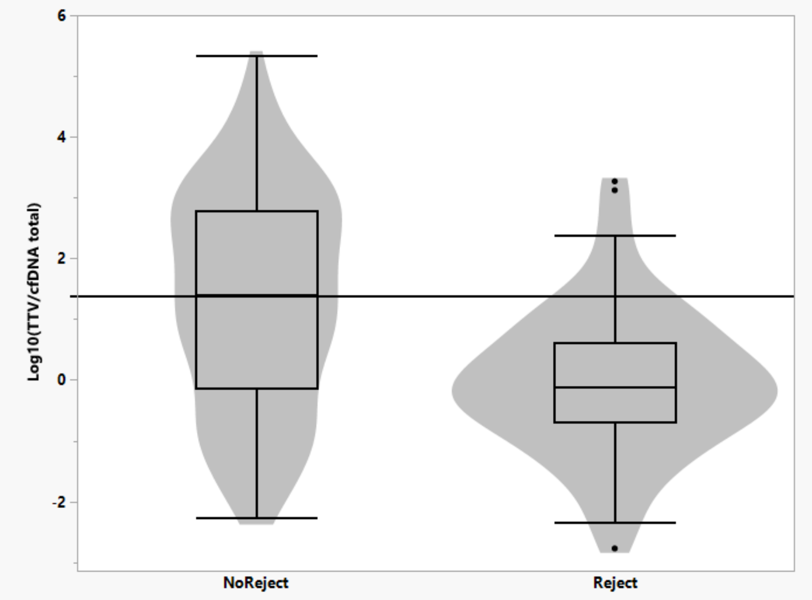


#### Oneway Analysis of Log10(TTV/cfDNA total) By Reject/NoReject

##### Quantiles

| Level | Minimum | 10% | 25% | Median | 75% | 90% | Maximum |
|---|---|---|---|---|---|---|---|
| NoReject | -2.28349 | -1.18796 | -0.14538 | 1.384554 | 2.776671 | 3.49844 | 5.3367 |
| Reject | -2.77124 | -1.53361 | -0.69742 | -0.12398 | 0.613755 | 1.399875 | 3.269291 |

##### t Test

Reject-NoReject

Assuming unequal variances

| | | | |
|---|---|---|---|
| Difference | -1.3447 | t Ratio | -6.85996 |
| Std Err Dif | 0.1960 | DF | 168.153 |
| Upper CL Dif | -0.9577 | Prob > \|t\| | <.0001* |
| Lower CL Dif | -1.7317 | Prob > t | 1.0000 |
| Confidence | 0.95 | Prob < t | <.0001* |

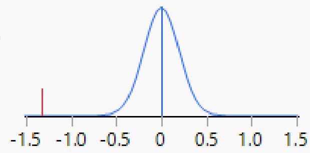


##### Wilcoxon / Kruskal-Wallis Tests (Rank Sums)

| Level | Count | Score Sum | Expected Score | Score Mean | (Mean-Mean0)/Std0 |
|---|---|---|---|---|---|
| NoReject | 192 | 27667.0 | 24864.0 | 144.099 | 5.359 |
| Reject | 66 | 5744.00 | 8547.00 | 87.030 | -5.359 |

###### Wilcoxon Two-Sample Test, Normal Approximation

| S | Z | Prob>Z | Prob>\|Z\| |
|---|---|---|---|
| 5744 | -5.35876 | <.0001* | <.0001* |

###### Kruskal-Wallis Test, ChiSquare Approximation

| ChiSquare | DF | Prob>ChiSq |
|---|---|---|
| 28.7266 | 1 | <.0001* |

### Figure S5. Delta Cq(VIC-FAM) and different biopsy outcomes

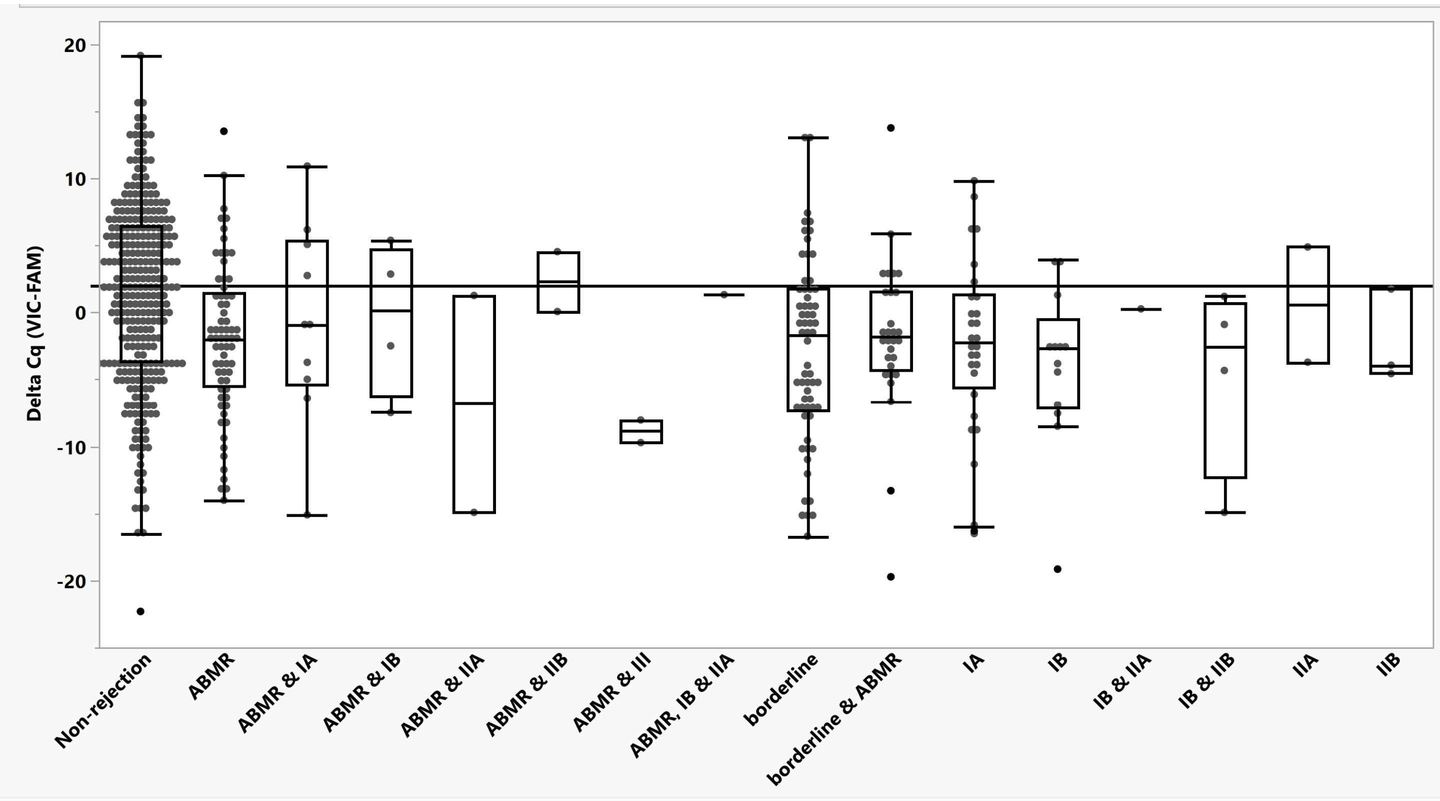

### Figure S6. Matched pairs test for sequential biopsy results

A.

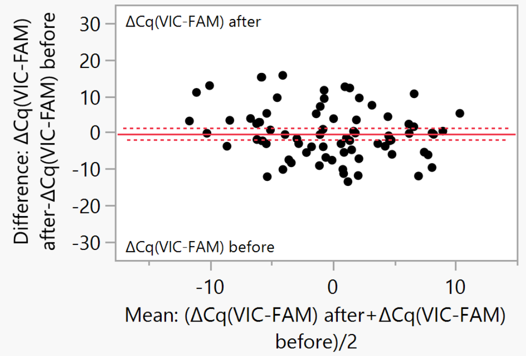


| | | | |
|---|---|---|---|
| ΔCq(VIC-FAM) after | -0.2951 | t-Ratio | -0.41023 |
| ΔCq(VIC-FAM) before | 0.04318 | DF | 72 |
| Mean Difference | -0.3383 | Prob > \|t\| | 0.6829 |
| Std Error | 0.82459 | Prob > t | 0.6586 |
| Upper 95% | 1.30552 | Prob < t | 0.3414 |
| Lower 95% | -1.9821 | | |
| N | 73 | | |
| Correlation | 0.38082 | | |

**Wilcoxon Signed Rank**

| | **ΔCq(VIC-FAM) after-ΔCq(VIC-FAM) before** |
|---|---|
| Test Statistic S | -133.50 |
| Prob>\|S\| | 0.4668 |
| Prob>S | 0.7666 |
| Prob<S | 0.2334 |

B.

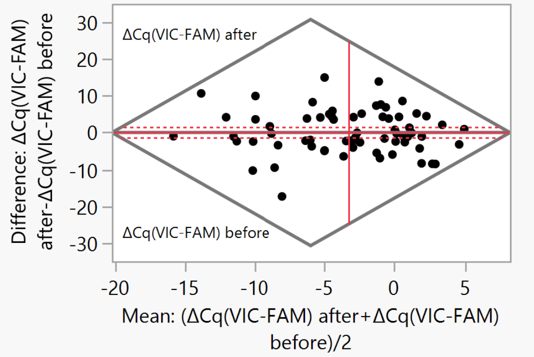


| | | | |
|---|---|---|---|
| ΔCq(VIC-FAM) after | -3.1853 | t-Ratio | 0.259595 |
| ΔCq(VIC-FAM) before | -3.3721 | DF | 66 |
| Mean Difference | 0.18679 | Prob > \|t\| | 0.7960 |
| Std Error | 0.71954 | Prob > t | 0.3980 |
| Upper 95% | 1.62339 | Prob < t | 0.6020 |
| Lower 95% | -1.2498 | | |
| N | 67 | | |
| Correlation | 0.4361 | | |

**Wilcoxon Signed Rank**

| | **ΔCq(VIC-FAM) after-ΔCq(VIC-FAM) before** |
|---|---|
| Test Statistic S | 30.000 |
| Prob>\|S\| | 0.8530 |
| Prob>S | 0.4265 |
| Prob<S | 0.5735 |

C.

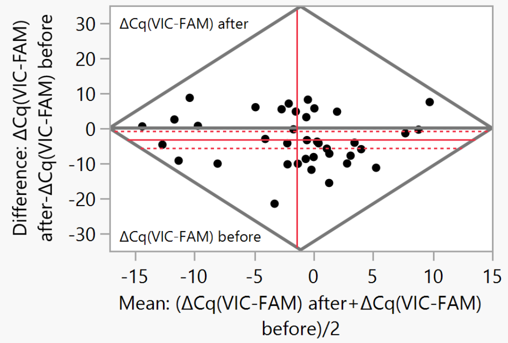


| | | | |
|---|---|---|---|
| ΔCq(VIC-FAM) after | -3.0335 | t-Ratio | -2.69388 |
| ΔCq(VIC-FAM) before | 0.13659 | DF | 37 |
| Mean Difference | -3.1701 | Prob > \|t\| | 0.0106* |
| Std Error | 1.17678 | Prob > t | 0.9947 |
| Upper 95% | -0.7857 | Prob < t | 0.0053* |
| Lower 95% | -5.5545 | | |
| N | 38 | | |
| Correlation | 0.42936 | | |

**Wilcoxon Signed Rank**

| | **ΔCq(VIC-FAM) after-ΔCq(VIC-FAM) before** |
|---|---|
| Test Statistic S | -165.50 |
| Prob>\|S\| | 0.0143* |
| Prob>S | 0.9929 |
| Prob<S | 0.0071* |

D.

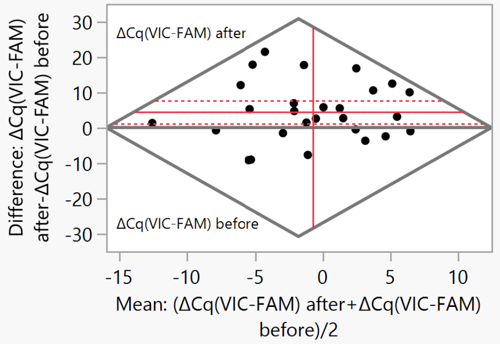


| | | | |
|---|---|---|---|
| ΔCq(VIC-FAM) after | 1.5319 | t-Ratio | 2.839746 |
| ΔCq(VIC-FAM) before | -2.9773 | DF | 26 |
| Mean Difference | 4.50921 | Prob > \|t\| | 0.0087* |
| Std Error | 1.58789 | Prob > t | 0.0043* |
| Upper 95% | 7.77317 | Prob < t | 0.9957 |
| Lower 95% | 1.24525 | | |
| N | 27 | | |
| Correlation | 0.15156 | | |

**Wilcoxon Signed Rank**

| | **ΔCq(VIC-FAM) after-ΔCq(VIC-FAM) before** |
|---|---|
| Test Statistic S | 105.000 |
| Prob>\|S\| | 0.0088* |
| Prob>S | 0.0044* |
| Prob<S | 0.9956 |

### Figure S7. BKV qPCR data – linearity assessment

A.

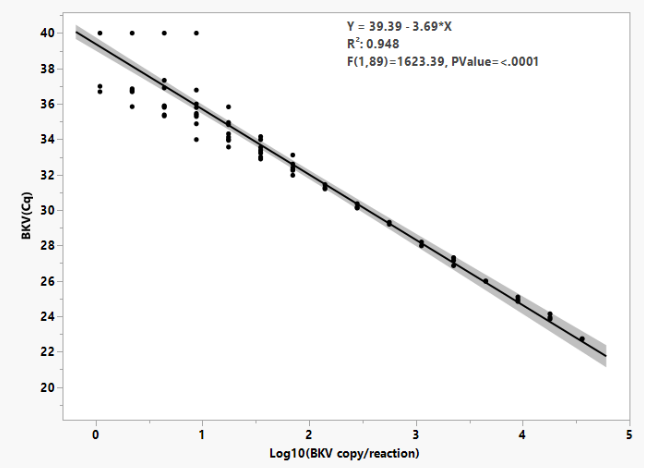


B.

| DNA input (copy/reaction) | Poisson distribution estimates for Cq(BKV) | | | |
|---|---|---|---|---|
| | λ | Std Error | Lower 95% CI | Upper 95% CI |
| 1 | 39.4 | 2.0 | 35.6 | 43.4 |
| 2 | 38.4 | 2.0 | 34.7 | 42.4 |
| 4 | 37.2 | 1.9 | 33.5 | 41.1 |
| 9 | 35.8 | 1.9 | 32.2 | 39.6 |
| 18 | 34.5 | 1.9 | 31.0 | 38.3 |
| 35 | 33.4 | 2.0 | 29.5 | 37.5 |
| 70 | 32.3 | 2.3 | 28.0 | 37.1 |

C.

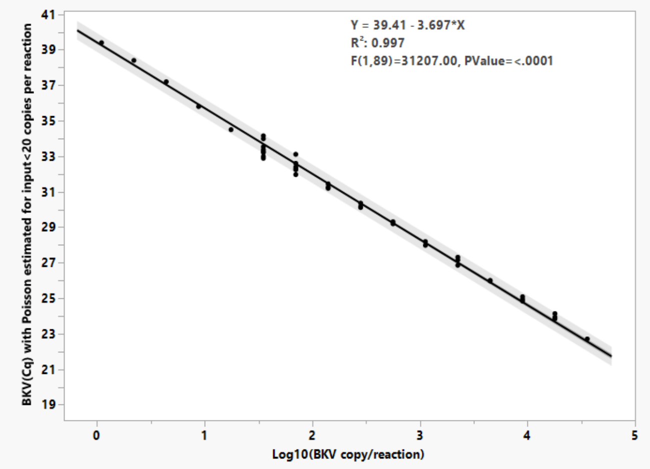


D.

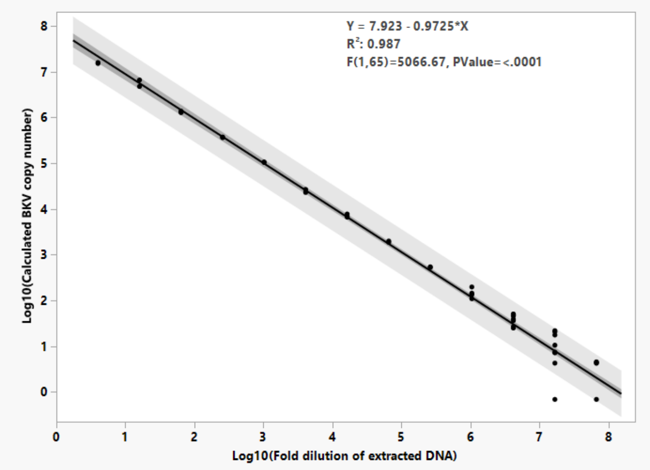

### Figure S8. BKV qPCR data and biopsy results

A.

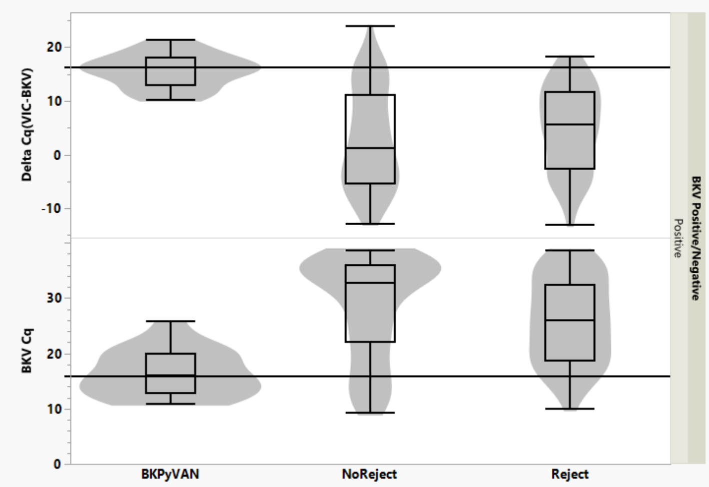


B.

**Quantiles**

| Level | Minimum | 10% | 25% | Median | 75% | 90% | Maximum |
|---|---|---|---|---|---|---|---|
| BKPyVAN | 10.09349 | 10.75815 | 12.87332 | 16.25235 | 18.02739 | 20.24796 | 21.29518 |
| NoReject | -12.8422 | -8.11726 | -5.40097 | 1.213976 | 11.08644 | 16.4196 | 23.85865 |
| Reject | -13.0789 | -5.90514 | -2.58881 | 5.594934 | 11.7305 | 14.52503 | 18.19941 |

**Means Comparisons**

**Comparisons with a control using Dunnett's Method**

Control Group = BKPyVAN

**LSD Threshold Matrix**

| Level | Abs(Dif)-LSD | p-Value |
|---|---|---|
| BKPyVAN | -6.3 | 1.0000 |
| Reject | 5.627 | <.0001* |
| NoReject | 8.001 | <.0001* |

Positive values show pairs of means that are significantly different.

C.

**Quantiles**

| Level | Minimum | 10% | 25% | Median | 75% | 90% | Maximum |
|---|---|---|---|---|---|---|---|
| BKPyVAN | 10.78228 | 11.06225 | 12.79879 | 15.91166 | 19.98235 | 22.11808 | 25.74903 |
| NoReject | 9.24703 | 12.78707 | 22.08395 | 32.76296 | 35.89028 | 36.9521 | 38.59869 |
| Reject | 9.909822 | 15.79212 | 18.75498 | 25.92817 | 32.414 | 36.70699 | 38.56838 |

**Means Comparisons**

**Comparisons with a control using Dunnett's Method**

Control Group = BKPyVAN

**LSD Threshold Matrix**

| Level | Abs(Dif)-LSD | p-Value |
|---|---|---|
| NoReject | 7.515 | <.0001* |
| Reject | 4.554 | <.0001* |
| BKPyVAN | -6.17 | 1.0000 |

Figure S9. Analysis of BKV positive samples, TTV, cfDNA and BKV quantification results

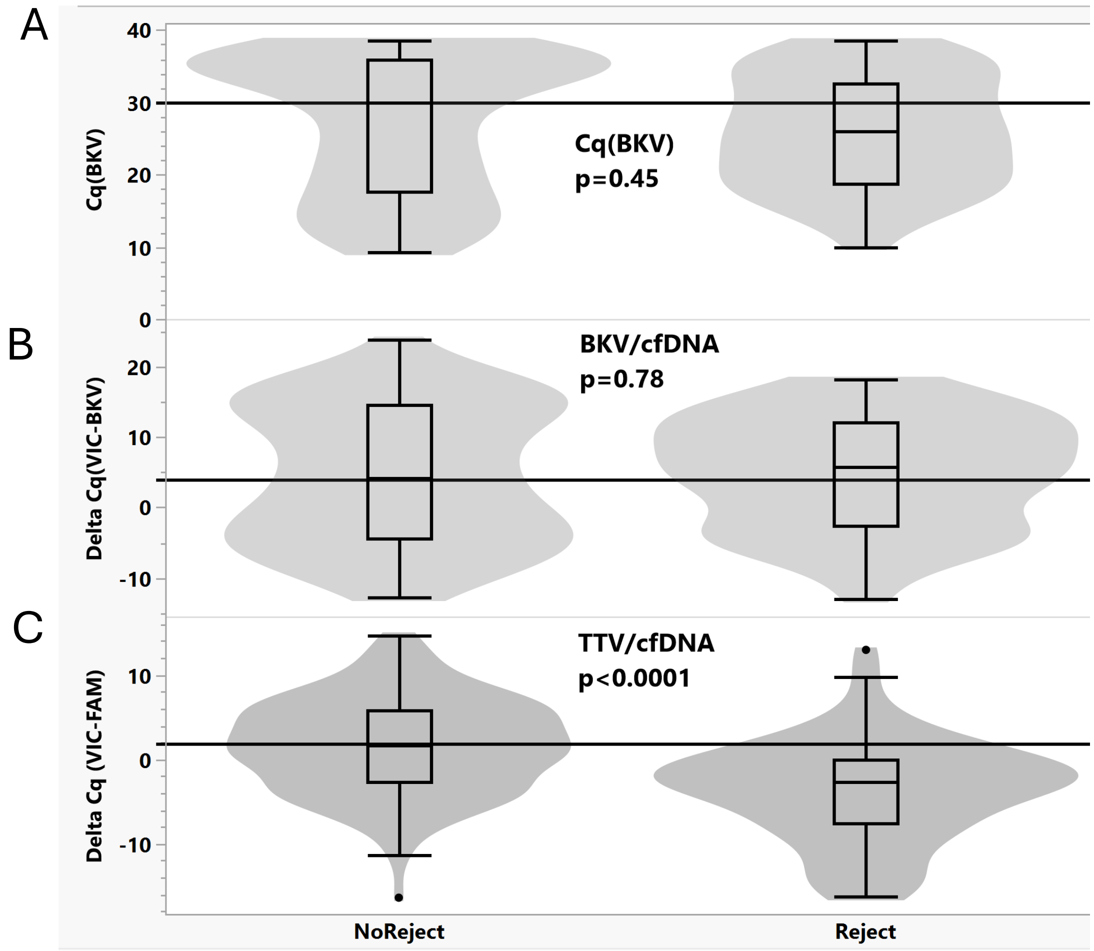


D

| Source | Logworth | PValue |
|---|---|---|
| Days post transplant | 2.387 | 0.00410 |
| RACE | 1.323 | 0.04749 |
| SEX | 1.293 | 0.05092 |
| BKV Cq | 0.812 | 0.15419 |
| Age at Time of Transplant | 0.098 | 0.79710 |

### Figure S10. Combined nominal logistic model for kidney rejection

A

| Source | Logworth | PValue |
|---|---|---|
| Delta Cq (VIC-FAM) | 4.894 | 0.00001 |
| RACE | 1.059 | 0.08738 |
| Days post transplant | 0.965 | 0.10828 |
| Total cfDNA (VIC Cq) | 0.723 | 0.18910 |
| SEX | 0.648 | 0.22512 |
| BKV Cq | 0.173 | 0.67137 |
| Age at Time of Transplant | 0.072 | 0.84648 |
| Delta Cq(VIC-JUN) | 0.058 | 0.87559 |

B

**For Reject/NoReject odds of Reject versus NoReject**

**Unit Odds Ratios**

**Per unit change in regressor**

| Term | Odds Ratio | Lower 95% | Upper 95% | Reciprocal |
|---|---|---|---|---|
| Delta Cq (VIC-FAM) | 0.865868 | 0.80421 | 0.925977 | 1.1549105 |
| Delta Cq(VIC-JUN) | 0.985753 | 0.833352 | 1.200098 | 1.0144524 |
| Total cfDNA (VIC Cq) | 0.936089 | 0.844141 | 1.032444 | 1.068275 |
| Age at Time of Transplant | 1.002444 | 0.977998 | 1.027857 | 0.9975618 |
| Days post transplant | 1.002331 | 0.999486 | 1.005243 | 0.997674 |
| BKV Cq | 0.990912 | 0.949589 | 1.033715 | 1.0091715 |

C

**Receiver Operating Characteristic**

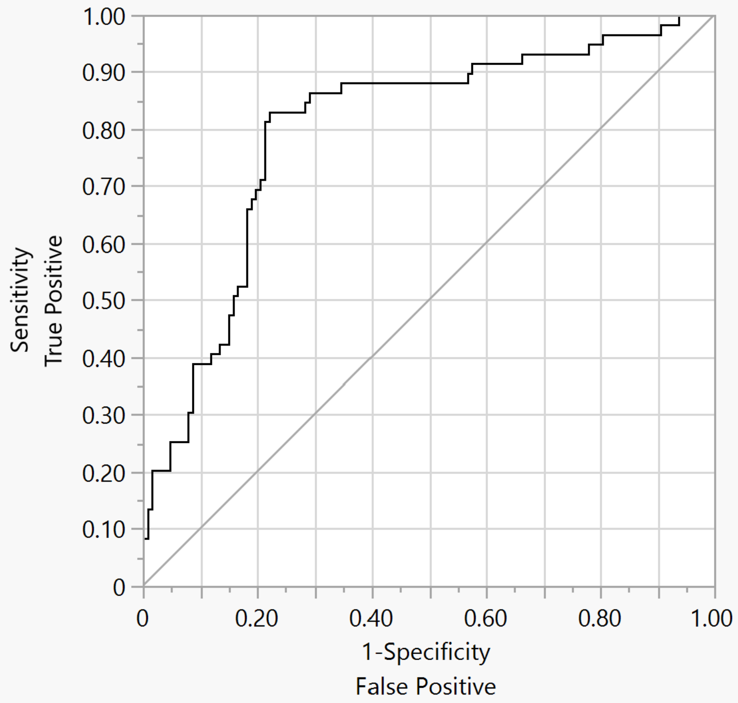


Using Reject/NoReject='Reject' to be the positive level

| AUC |
|---|
| 0.80061 |

### Figure S11. Nominal logistic model for different types of rejection

A

| Source | Logworth | PValue |
|---|---|---|
| Delta Cq (VIC-FAM) | 3.592 | 0.00026 |
| Days post transplant | 1.940 | 0.01147 |
| SEX | 1.029 | 0.09344 |
| RACE | 0.592 | 0.25577 |
| Delta Cq(VIC-JUN) | 0.374 | 0.42245 |
| Total cfDNA (VIC Cq) | 0.206 | 0.62177 |
| Age at Time of Transplant | 0.188 | 0.64797 |
| BKV Cq | 0.029 | 0.93542 |

B

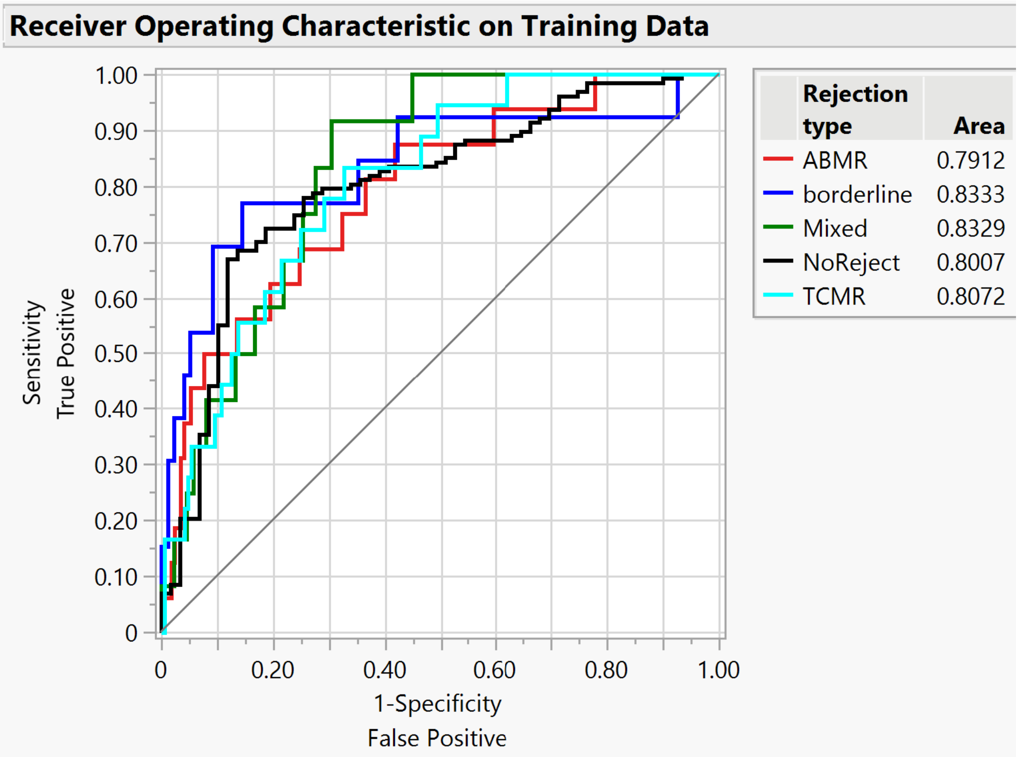

### Figure S12. Comparing Cq(BKV) and delta Cq(VIC-BKV) models

A

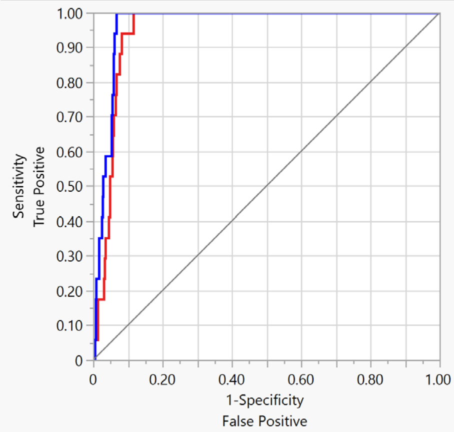


| Predictor | Creator | AUC |
|---|---|---|
| Prob[BKPyVAN] Cq(BKV) | Fit Nominal Logistic | 0.9509 |
| Prob[BKPyVAN] Delta Cq(VIC-BKV) | Fit Nominal Logistic | 0.9667 |

B

| Predictor | AUC | Std Error | Lower 95% | Upper 95% |
|---|---|---|---|---|
| Prob[BKPyVAN] Cq(BKV) | 0.9509 | 0.0105 | 0.9258 | 0.9678 |
| Prob[BKPyVAN] Delta Cq(VIC-BKV) | 0.9667 | 0.0083 | 0.9459 | 0.9796 |

| Predictor | vs. Predictor | AUC Difference | Std Error | Lower 95% | Upper 95% | ChiSquare | Prob>ChiSq |
|---|---|---|---|---|---|---|---|
| Prob[BKPyVAN] Cq(BKV) | Prob[BKPyVAN] Delta Cq(VIC-BKV) | -0.016 | 0.0079 | -0.031 | -0.000 | 4.002 | 0.0454* |

| Test | ChiSquare | DF | Prob>ChiSq |
|---|---|---|---|
| All AUCs equal | 4.00197 | 1 | 0.0454* |

C

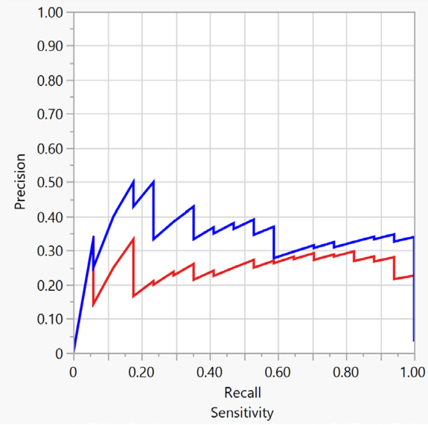


| Predictor | Creator | Precision-Recall AUC |
|---|---|---|
| Prob[BKPyVAN] Cq(BKV) | Fit Nominal Logistic | 0.2467 |
| Prob[BKPyVAN] Delta Cq(VIC-BKV) | Fit Nominal Logistic | 0.3446 |

D

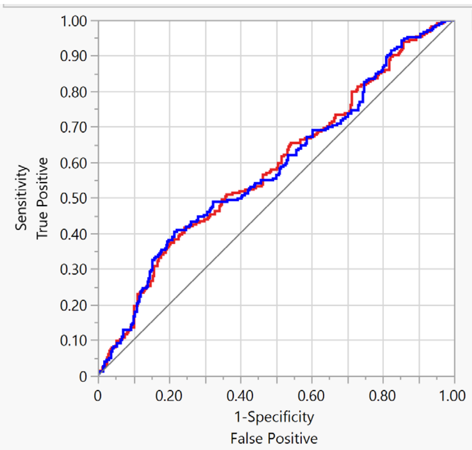


| Predictor | Creator | AUC |
|---|---|---|
| Prob[Reject] Cq(BKV) | Fit Nominal Logistic | 0.5843 |
| Prob[Reject] Delta Cq(VIC-BKV) | Fit Nominal Logistic | 0.5861 |
