## Supplemental tables for "To TTV or Not to TTV? A Combined Urinary Biomarker Approach in Kidney Transplant Recipients Using Torque Teno Virus, Cell-Free DNA, and BK Polyomavirus Measurements"

**Table S1. Sample information summary for initial case-control study**

| Parameter | Sample Information[a] | |
|---|---|---|
| Age; years, median (quartiles) | 52 | (39 — 63) |
| Age; years, min — max (range) | 20 — 79 | (59) |
| Sex; male / female, n (%) | 176 / 105 | (62.6 / 37.4) |
| Race; n (%) | | |
| Asian | 20 | (7.1) |
| Black | 52 | (18.5) |
| White | 204 | (72.6) |
| Pacific Islander | 2 | (0.7) |
| Not provided | 3 | (1.1) |
| Biopsy Type, n (%) | | |
| Protocol | 195 | (69) |
| For-cause | 86 | (31) |
| Days post transplant, median (quartiles) | 119 | (90 — 346) |
| Days post transplant, min — max (range) | 4 — 685 | (681) |
| Biopsy results, n (%) | | |
| No Rejection | 117 | (41.6) |
| TCMR | 32 | (11.4) |
| Borderline | 42 | (14.9) |
| Mixed | 40 | (14.2) |
| ABMR | 50 | (17.8) |
| All Types of Rejection | 164 | (58.4) |

[a] information is summarized for 281 samples obtained from 158 kidney transplant patients

**Table S2. Sample information for confirmatory study**

| Parameter | Sample Information[a] | |
|---|---|---|
| Age; years, median (quartiles) | 53 | (40 — 62) |
| Age; years, min — max (range) | 18 — 79 | (61) |
| Sex; male / female, n (%) | 203 / 97 | (67.7 / 32.3) |
| Race; n (%) | | |
| Asian | 13 | (4.3) |
| Black | 60 | (20.0) |
| White | 214 | (71.3) |
| Pacific Islander | 2 | (0.7) |
| Not provided | 11 | (3.7) |
| Biopsy Type, n (%) | | |
| Protocol | 217 | (72.3) |
| For-cause | 83 | (27.7) |

| | | |
|---|---|---|
| Days post transplant, median (quartiles) | 106 | (87 — 256) |
| Days post transplant, min — max (range) | 14 — 613 | (599) |
| Biopsy results, n (%) | | |
| No Rejection | 193 | (64.3) |
| TCMR | 23 | (7.7) |
| Borderline | 31 | (10.3) |
| Mixed | 19 | (6.3) |
| ABMR | 34 | (11.3) |
| All Types of Rejection | 107 | (35.7) |

[a] information is summarized for 300 samples obtained from 240 kidney transplant patients

**Table S3. Models for kidney rejection generated for the initial set of 281 samples and the performance of resulting probability formulas in the confirmatory data set.**

| Type of analysis | First study (281 samples) | Second study (300 samples) |
|---|---|---|
| Comparing ΔCq(VIC-FAM) in rejection and no rejection groups | | |
| T-test, p-value | 0.0026 | <0.0001 |
| Wilcoxon test, p-value | 0.0015 | <0.0001 |
| Model performance comparison between training and confirmatory data sets (AUROC, 95% CI) | | |
| Generalized linear model for all types of rejection[a] | 0.64 (0.57 - 0.71) | 0.73 (0.67-0.78) |
| Boosted neural model for all types of rejection[a] | 0.68 (0.61 – 0.74) | 0.72 (0.65 - 0.77) |
| Generalized linear model for ABMR | 0.70 (0.61 – 0.78) | 0.71 (0.63 – 0.79) |
| Generalized linear model for TCMR | 0.69 (0.57 – 0.75) | 0.69 (0.58 – 0.79) |

[a] Model was generated using qPCR data, age of the patient at the time of transplant and number of days between transplant and sample collection. Probability formula created with first set of 281 samples was applied to data from the second study. AUROC values for second subset of samples were calculated using probability formula generated with 281 samples in the first study.

**Table S4. Subset of samples used for digital PCR based analysis**

| Parameter | Sample Information[a] | |
|---|---|---|
| Age; years, median (quartiles) | 52 | (40 — 62) |
| Age; years, min — max (range) | 18 — 79 | (61) |
| Sex; male / female, n (%) | 181 / 77 | (70.2 / 29.8) |
| Race; n (%) | | |
| Asian | 11 | (4.3) |

| | | |
|---|---|---|
| Black | 67 | (26.0) |
| White | 170 | (65.9) |
| Pacific Islander | 2 | (0.8) |
| Not provided | 8 | (3.1) |
| Biopsy Type, n (%) | | |
| Protocol | 194 | (75.2) |
| For-cause | 64 | (24.8) |
| Days post transplant, median (quartiles) | 106 | (89 — 253) |
| Days post transplant, min — max (range) | 14 — 613 | (599) |
| Biopsy results, n (%) | | |
| No Rejection | 192 | (74.4) |
| TCMR | 12 | (4.7) |
| Borderline | 21 | (8.1) |
| Mixed | 8 | (3.1) |
| ABMR | 25 | (9.7) |
| All Types of Rejection | 66 | (25.6) |

[a] Subset of 258 samples randomly selected from confirmatory group of 300 samples initially tested by qPCR

**Table S5. Analysis of ΔCq(VIC-FAM) for different types of rejection**

| Type of rejection | N | Mean (95% CI, Lower/Upper) | P-value in t-test [a] | P-value in Wilcoxon signed rank test [a] |
|---|---|---|---|---|
| No Rejection | 312 | 1.47 (0.72 / 2.21) | 0.0110 | 0.0026 |
| Borderline | 63 | -2.87 (-4.58 / -1.15) | 0.0002 | 0.0002 |
| TCMR | 51 | -2.77 (-4.47 / -1.08) | 0.0003 | 0.0002 |
| ABMR | 68 | -1.99 (-3.37 / -0.61) | 0.0006 | 0.0004 |
| Mixed | 50 | -1.73 (-3.50 / 0.03) | 0.0142 | 0.0108 |

[a] One sample t-test and non-parametric Wilcoxon signed rank test were performed with 0.5 ΔCq selected as a potential cutoff. Two-sided p-values are shown.

**Table S6. Subset of samples used for BKV analysis**

| Parameter | Sample Information | |
|---|---|---|
| Age; years, median (quartiles) | 53 | (40 — 63) |
| Age; years, min — max (range) | 18 — 79 | (61) |
| Sex; male / female, n (%) | 337 / 186 | (64.4 / 35.6) |
| Race; n (%) | | |
| Asian | 25 | (4.8) |
| Black | 108 | (20.7) |
| White | 374 | (71.5) |
| Pacific Islander | 3 | (0.6) |
| Not provided | 13 | (2.5) |

| | | |
|---|---|---|
| Biopsy Type, n (%) | | |
| Protocol | 376 | (71.9) |
| For-cause | 147 | (28.1) |
| Days post transplant, median (quartiles) | 109 | (88 — 280) |
| Days post transplant, min — max (range) | 4 — 685 | (681) |
| Biopsy results, n (%) | | |
| No Rejection | 304 | (58.1) |
| All Types of Rejection | 219 | (41.9) |
| Biopsy results for BKV associated polyoma, n (%) | | |
| BK Polyoma detected/no rejection | 17 | (3.3) |
| BK Polyoma detected/rejection detected | 5 | (1.0) |
| No BK polyoma | 501 | (95.8) |
| Biopsy results, rejection type, n (%) | | |
| No rejection | 304 | (58.1) |
| ABMR | 65 | (12.4) |
| Borderline | 59 | (11.3) |
| TCMR | 46 | (8.8) |
| Mixed rejection | 49 | (9.4) |
